# SIENNA: Lightweight Generalizable Machine Learning Platform for Brain Tumor Diagnostics

**DOI:** 10.1101/2024.04.03.24305210

**Authors:** Sreya Sunil, Rahul S. Rajeev, Ayan Chatterjee, Julie Pilitsis, Amitava Mukherjee, Janet L. Paluh

## Abstract

The transformative integration of Machine Learning (ML) for Artificial General Intelligence (AGI)-enhanced clinical imaging diagnostics, is itself in development. In brain tumor pathologies, magnetic resonance imaging (MRI) is a critical step that impacts the decision for invasive surgery, yet expert MRI tumor typing is inconsistent and misdiagnosis can reach levels as high as 85%. Current state-of-the-art (SOTA) ML brain tumor models struggle with data overfitting and susceptibility to shortcut learning, further exacerbated in large-sized models with many tunable parameters. In a comparison with multiple SOTA models, our deep ML brain tumor diagnostics model, SIENNA, surpassed limitations in four key areas of prioritized minimal data preprocessing, an optimized architecture that reduces shortcut learning and overfitting, integrated inductive cross-validation method for generalizability, and smaller neural architecture. SIENNA is applicable across MRI machines and 1.5 and 3.0 Tesla, and achieves high average accuracies on clinical DICOM MRI data across three-way classification: 92% (non-tumor), 91% (GBM), and 93% (MET) with retained high F1 and AUROC values for limited false positives/negatives. SIENNA is a lightweight clinical-ready AGI framework compatible with future multimodal expanded data integration.

## INTRODUCTION

Artificial intelligence (AI) in medical imaging diagnostics is poised to revolutionize patient care in the coming decades yet grapples with the challenge of handling data while preserving physiological complexity. This impact will be especially pronounced in the clinical diagnosis, monitoring, and treatment of brain tumor neoplasms. Brain tumors, the most common malignancy of the central nervous system (CNS), rank 10th globally among leading causes of death [1,2]. The clinical multi-classification of tumors heavily relies on diagnostic expertise, particularly utilizing MRI [4]. Since its clinical introduction for brain tumor detection in 1985 [3,4], MRI usage has surged, with roughly 34 machines per million people in the United States alone by 2020, resulting in over 30 million images captured [5,6]. However, accurately recognizing morphological features remains challenging [7]. CNS tumor types and grades exhibit variability influenced by factors such as sex, age, and demographics [2,8,9], contributing to an alarming 85% misdiagnosis rate for prominent brain tumor types via MRI, as revealed by retrospective analyses [10]. This study highlights the challenges with obtaining uniform high accuracy in diagnoses across clinicians and centers, which will benefit from AI technologies. The intricate variability in data across pathologies and patients further underscores the necessity for new AI platforms and the need to exercise particular care in data handling, advancing image comparability algorithms, and limiting data preprocessing. Relying on highly preprocessed public datasets creates challenges for generalizability of the AI platforms for advancing towards AGI [11]. Furthermore, shortcut learning [12,13] has recently emerged as a significant drawback in various ML approaches, alongside the well-known issue of overfitting [14]. In shortcut learning, ML models struggle to capture the desired morphological features of images and instead resort to exploiting undesired patterns to achieve high cross-validation performance. For instance, Geirhos et al. [12] demonstrated that a ML model trained to identify pneumonia from lung scans achieves high performance by capitalizing on hospital tokens within the images rather than learning the morphological patterns, rendering it ineffective in real-world scenarios. Consequently, when deployed in practical settings, these models struggle to generalize to new, unseen data [15]. Moreover, larger models with numerous parameters are particularly susceptible to overfitting and shortcut learning. Recent studies emphasize the importance of smaller ML models with fewer parameters for enhanced generalization [16], a crucial step in the development of AGI [17]. Furthermore, traditional cross-validation techniques merge image data for evaluating performance irrespective of patient information. Thus, this form of test performance if misleading in the clinical setting when the model encounters images from a new patient.

For AI design and data handling to achieve greatest impact clinically and to be increasingly patient-informative the focus on generalizable platforms that can retrieve and learn from new diverse morphological information in clinical data is essential [7]. Retention of physiological detail in data handling will determine the extent to which AI can link mechanistic information to near imperceptible tissue variations. Metastasized secondary brain neoplasms (METs) arise from non-brain primary cancers [10], whereas glioblastomas (GBMs) are prominent intracranial in origin tumors [1,2,11,18]. A multitude of primary cancer sites that contribute to brain METs include the lung (39-56 percent), breast (13-30 percent), melanomas (8-11 percent), gastrointestinal (6-9 percent) and renal (2-6 percent) tissues [10]. To what extent METs reflect aspects of the primary cancer tissue source is still being studied, but correlative data on brain tumor morphology and distribution patterns suggest some physiological links [10, 19-22]. The underlying genetic signatures of GBMs may also impact physiologic heterogeneity and impact current survival beyond 5 years diagnosis [11,23]. Data handling in AGI platforms will be key to lay the framework for future AI subtyping studies with potential to expand diagnostics and treatments. The current challenges in expert multi-classification of normal physiology, GBM, or MET tumors [9,24] are reflected in the reliance on a range of additional clinical information [25] that includes a) patient history of systemic malignancy, tumor features, cerebral location, positioning near gray or white matter, a multiplicity of cerebral sites [26], b) MR tumor morphology [27], c) MR perfusion for tumor vascularity [28,29], d) use of multiple MRI modalities [30] and e) intraoperative biopsies for biomarker typing of tumors [31]. The overwhelming and growing need for MRI diagnostics with expanded associated technology is expected to continue to strain clinical diagnostic accuracy. Both 1.5 and 3.0 Tesla MRI images are currently dominant in clinical practice [32]. However 7.0 Tesla MRI is approved for use clinically [33] and includes physiological detail beyond the training of most clinicians. Multiparameter MRI [34] and portable lower Tesla spectrums [35] are also increasing demand for clinical AGI in MRI [36, 37].

An emerging barrier to advancing clinical AGI platforms for MRI diagnostics is non-optimal data preprocessing. AI models that employ public preprocessed datasets, such as the BraTS 2018 and BraTS 2020 MRI datasets [38,39], exclusively in their development run the risk of being compromised for generalizability. Misprioritization of features will reduce accuracy when moving from public to minimally processed clinical datasets of Digital Imaging and Communications in Medicine (DICOM) origin [40] that is attributable to a gap in ML platforms between design and data preprocessing. Complex preprocessing operations such as skull stripping [41] and MR bias field correction [40] can increase initial accuracy by eliminating noise but are suboptimal for clinical data diagnostics. Thus, while some studies on multi-classification models may seem promising as diagnostics across tumor types [43] or for subtypes of tumors, [44,45] their training on highly processed datasets will limit their generalizability and transferability to new patients outside the training set in a clinical setting. Furthermore, traditional cross-validation on the benchmark datasets fails to report a model’s ability to generalize to new patients. We previously developed the SCENIC [46] convolutional neural network (CNN) architecture for neoplasm identification and achieved 98.3% classification accuracy on the publicly available BraTS Dataset. SCENIC outperforms and is competitive versus multiple other tested approaches that include XceptionNet [47], InceptionV3 [48], ResNet-50 [49], and VGG-16 [50]. Although SOTA for identifying tumor presence across MRI modalities, SCENIC design is not immediately generalizable to high accuracy multi-classification of minimally processed clinical data for GBM and MET tumors. Achieving data analysis generalizability along with robust accuracy is of high priority to expand the applications possible for MRI AI architectures and their ability to meet clinical diagnostic challenges and keep pace with advancing MRI technologies.

In recent literature, various ML models have been developed to address different facets of brain tumor classification. NeuroXAI [51], for instance, integrates seven explanation methods, including attention-based explanations, to enhance the interpretability of ML predictions. Sturgeon’s approach [52] utilizes simulated nanopore sequencing data derived from readily available methylation array data, enabling accurate classification of tumor types based on intraoperatively generated sequence data. OpenSRH [53] employs optical histology data to train a self-supervised model for brain tumor diagnostics. VUNO Med-DeepBrain AD (DBAD) [54] leverages deep learning algorithms, serving as a decision support service for Alzheimer’s disease diagnosis. Additionally, Richardson et al. [55] conducted a genome-wide analysis of glioblastoma patients with unexpectedly long survival. Despite the proliferation of ML and non-ML approaches in brain tumor diagnostics, the literature often neglects to emphasize either generalizability or real-time deployability of these models in clinical settings. This oversight is particularly associated with the size and fine-tuning requirements of these models.

Here we introduce SIENNA, a light-weight multilayer CNN deep learning architecture with minimal preprocessing of clinical MR DICOM images. We demonstrate SIENNA’s generalizability to new patients, robustness, and high accuracy in clinical brain tumor diagnostics. SIENNA exhibits minimal misdiagnosis risk by use of a non-inter-dependent multi-classification approach that separately evaluates normal physiology and GBM and MET tumor pathophysiologies. SIENNA outperforms other ML technologies developed on highly processed public datasets in inducive tests. We identify and describe key challenges and strategies for retaining spatial and depth dimensional physiological features in working with data extraction from clinical MR DICOM images while reducing variability that interferes with comparability and can arise during data acquisition. This includes patient movement, scan acquisition parameters/conditions, and MRI Tesla levels, that impact image quality and scan intensities. SIENNA data handling includes the use of an in-house custom histogram equalization tool, PREMO, inspired by existing algorithms, [56]. PREMO applies pixel redistribution across intensity levels, enhancement of equalization by masking, and optimization by Gamma fine-tuning brightness and contrast. SIENNA’s ability to identify meaningful patterns and attributes is further enriched by adversarial training, [57,58] using images with subtle parameter distortion to exploit the model’s vulnerabilities and decision boundaries for re-optimization. Finally, we apply hyperparameter tuning [59,60] to SIENNA for robustness in diagnostic output, as has been demonstrated for CNN analysis applied in genetics [59], vision [61], histopathology [62] and with biomedical imaging [63]. SIENNA demonstrates on average accuracy on clinical DICOM MRI data across 3 tasks of 92% (Non-Tumor, SD=5.5%), 91% (GBM, SD = 3.2%), and 93% (MET, SD = 2.6%), with the distribution of accuracies skewed higher to 100% and a lower bound at 75%. SIENNA’s demonstrated generalizability and ability to outperform other AI platforms on clinical DICOM and publicly processed clinical data, is expected to accelerate companion diagnostic AI resources for clinical settings.

## RESULTS

### Co-development of SIENNA Deep Learning Architecture with Minimal Preprocessing of DICOM MRI data

We previously developed SCENIC [46] a deep learning architecture for robust tumor detection across MRI modalities of Fluid Attenuated Inversion Recovery (FLAIR), T1- and T2-weighted (T1-w; T2-w), and T1-contrast enhanced (T1-ce). SCENIC applied the BraTS 2017 dataset [38,39] of 1.5T MRI images from Gliomas, Meningioma and Pituitary tumors, or healthy tissue, achieving overall accuracy of 98.3 percent. Our de novo development of SIENNA ML architecture in this study was motivated by our inability to sufficiently generalize the SCENIC architecture to achieve similar high accuracy analysis of minimally processed MRI DICOM clinical images. The SIENNA AI architecture is developed along with strategies for minimal DICOM data preprocessing for comparability (**Fig1**) and avoids overprocessing of MRI data that is present in current public datasets and which limits generalizability to the clinic (**Fig.1a**) [64]. SIENNA incorporates a robust non-interdependent three-class multi-classification to discriminate between healthy tissue, or GBM or MET tumor pathology (**Fig.1b**). To reduce noise in our dataset with minimal processing we remove edge of scan outlier images of patient Z-stacks (**Fig.1c**). In **Fig.1d** we summarize features of data handling, including use of our custom histogram equalization software, PREMO, to correct images for variation in pixel intensity distribution from data acquisition, and adversarial training to challenge data feature learning, and ensure class balance. The SIENNA CNN architecture (**Fig.1e**) includes multiple feature extraction layers designed to map local and high-order spatial detail of the brain image, that are flattened and connected by dense/fully connected layers. The design enables the CNN network to learn and differentiate between different tumor types, resulting in a final layer that maps input to class probabilities. Shallow (inner) layers of the SIENNA CNN architecture are used to capture low-level features such as edges and local patterns of the brain, while the deeper layers extract more complex information such as global contrast relationships of the tumor. The performance of the native model is further optimized by tuning different combinations of hyperparameters for task-specific implementations. HYPERAS and the HYPEROPT library [49] are used to rapidly explore a range of user-defined search spaces, determined through data-driven experimentation and insights from prior literature [59-63]. Details are presented in Methods and the following specific analysis.

**Fig. 1.**
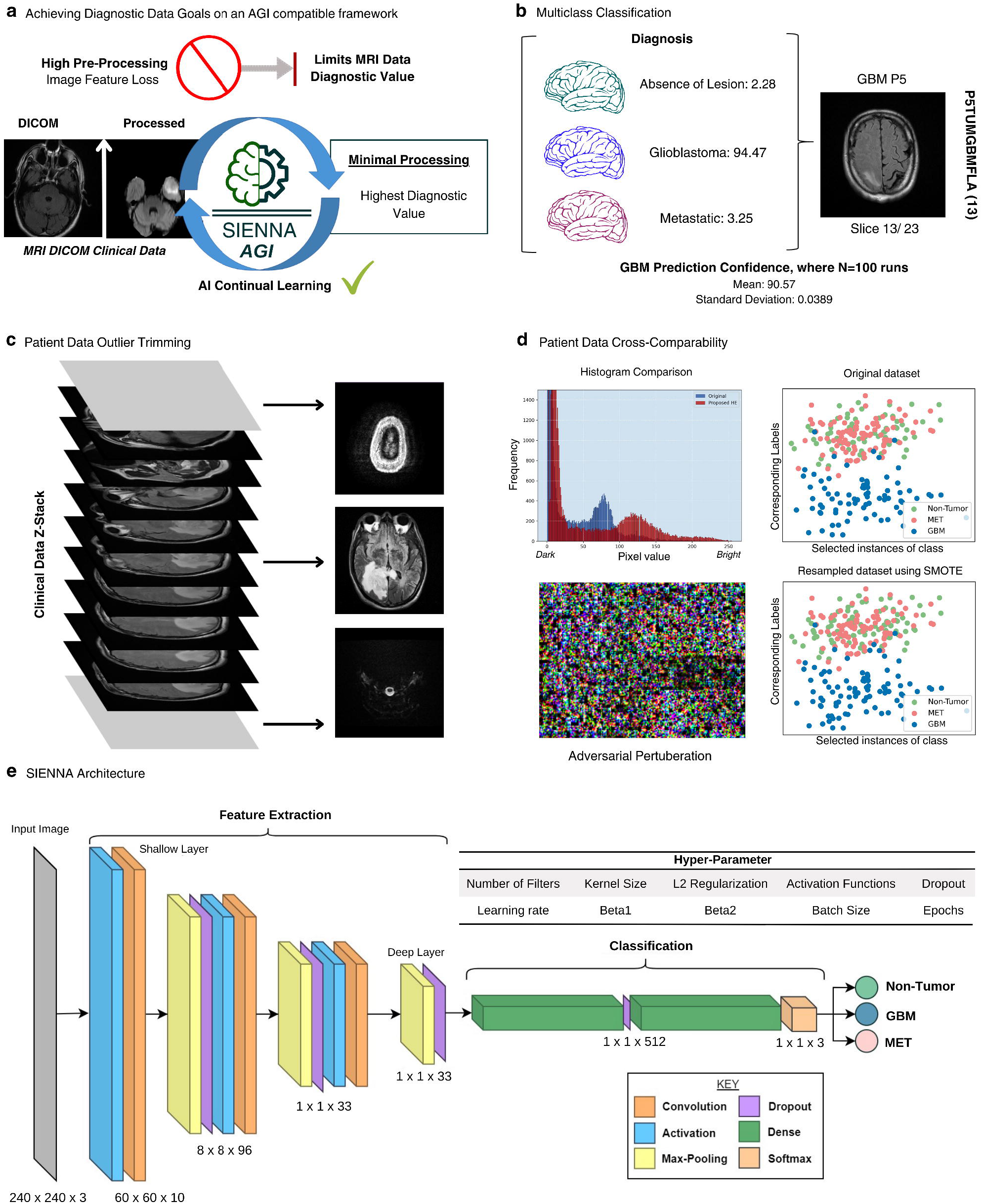
Overview of SIENNA Data Handling and CNN Deep Learning Architecture for Tumor Diagnostic Multi-classification. a. Clinical diagnostics requires generalizability that is lost by over-processing data and restricts future applications. b. SIENNA applies non-interdependent multi-classification to detect tumors, and GBM or MET types. The confidence percentage representing the level of certainty is obtained across these three classes for each 2D MRI scan of a dataset. Patient diagnostics is spatially detailed since each slice of the MRI data receives a probability assignment. c. Small dataset analysis benefits by removal of low information flanking Z-stacks. d. Data modification techniques utilized for model robustness. (top to bottom) Histogram plot comparison before and after equalization of pixel intensities and contrast sharpening using PREMO; adversarial perturbation with magnitude 0.1 imperceptible to human eyes. e. SIENNA architecture is a multilayer linear CNN deep learning model optimized via hyperparameter tuning. The architecture layer roles incorporate both spatial and depth dimensions and are color-coded.

### Multistep Preparation of De-identified Patient DICOM Data for Deep Learning Analysis

To address image quality and data comparability we generated a custom histogram equalizaiton platform, addressed class imbalance and class discrimination, and applied hyperparameter training for CNN optimization (**Fig.2** and **Methods**). We avoided commonly used extensive preprocessing techniques, including reorientation to LPS/RAI [65], skull-stripping [41], and N4 Bias correction [42], frequently applied in publicly available data. In our study, we assess a de-identified clinical sample database of 17 patient files (**Fig.3a**), including 9 GBM and 8 MET patients pre-validated by expert multifactorial clinical diagnostic standards [25] and obtained through IRB-approved collaboration (see **Methods**). We focus on MR fluid-attenuated inversion recovery (FLAIR) images, given the high dimensionality of that data and its ability in early testing to effectively distinguish tumor physiology in MR diagnostics [18]. To decrease the impact of noise interference in a small dataset, while retaining maximum information, we chose to remove a limited number of horizontal axial Z-slice scans, at the uppermost Z-scan that predominantly shows the skull area and lowermost scans containing primarily non-brain neck region tissues (**Fig.1c**). The final MRI DICOM clinical patient dataset comprised 386 scans, including 153 GBM and 125 MET clinically identified brain tumors, and 108 non-tumor scans.

SIENNA training, validation, and testing are done on shuffled individual Z-stacks from combined patient datasets, analyzing 386 patients’ Z-stack horizontal MRI scans. The variable sizes of brain tumors among the 17 patients’ data examined impact the number of tumor and non-tumor data images within a given patient horizontal Z-stack and GBM or MET tumor slices. A large tumor will constitute most horizontal slices in a patient dataset and the non-tumor scan becomes a minority class. To avoid class imbalance during training that can lead to biased models in small datasets that favor the majority class and fail to adequately learn the patterns and features of the minority class, we applied the Synthetic Minority Over-Sampling Technique (SMOTE) [66]. By SMOTE, synthetic samples are generated by interpolating between minority class examples (**Fig.1d** (scatter plot)). The MET and non-tumor classes were under-represented and deemed minority classes. For each minority instance chosen, its k (k=8 in our case) nearest neighbors are pinpointed based on Euclidean Distance. Interpolation then occurs between the feature vectors of this chosen instance and each of its k neighbors, resulting in the creation of synthetic instances. This procedure not only ensures that these newly minted samples align well with the original data’s distribution but also effectively balances the representation of minority classes. The scatter plot in **Fig.1d** provides a visual representation, demonstrating the equilibrium achieved between MET and GBM points before and after the application of SMOTE.

Comparability of data requires addressing variability arising during scan acquisition across different MRI systems including intensities and DICOM image quality across a diverse variety of patients and resources [41]. We conducted a comparative analysis of mulitple histogram equalization algorithms (**Fig.2a**), including Traditional Histogram Equalization [67], Adaptive Histogram Equalization [56], Contrast Histogram Equalization, and Contrast Limited Adaptive Histogram Equalization (CLAHE) [68] using the Structural Similarity Index (SSIM) [69]. These existing algorithms lacked the flexibility we desired to selectively apply enhancements based on specific regions of interest and resulted in off-target over-enhancement, loss of important details, and diminished diagnostic accuracy. To overcome these challenges, we generated a novel histogram equalization method that is inspired by existing algorithms but addresses brain regions of interest, Gamma correction, and other enhancements, that we refer to here as “Pixel Redistribution Enhancement, Masking, Optimization” or PREMO (see **Methods**). The PREMO algorithm uniformly redistributes pixel frequencies across intensity levels (**Fig. 2a** rightmost image), addressing the concentration of frequencies in dark pixels (**Fig. 2b**). It utilizes a binary mask to segment out non-brain regions and selectively apply enhancements. Furthermore, we introduced the adjustment of the Gamma value in PREMO to fine-tune brightness and contrast. This allowed for optimized visualization of subtle details and fine structures within the brain slices, enabling improved recognition of tumor-specific distinctive features by the SIENNA algorithm.

**Fig. 2.**
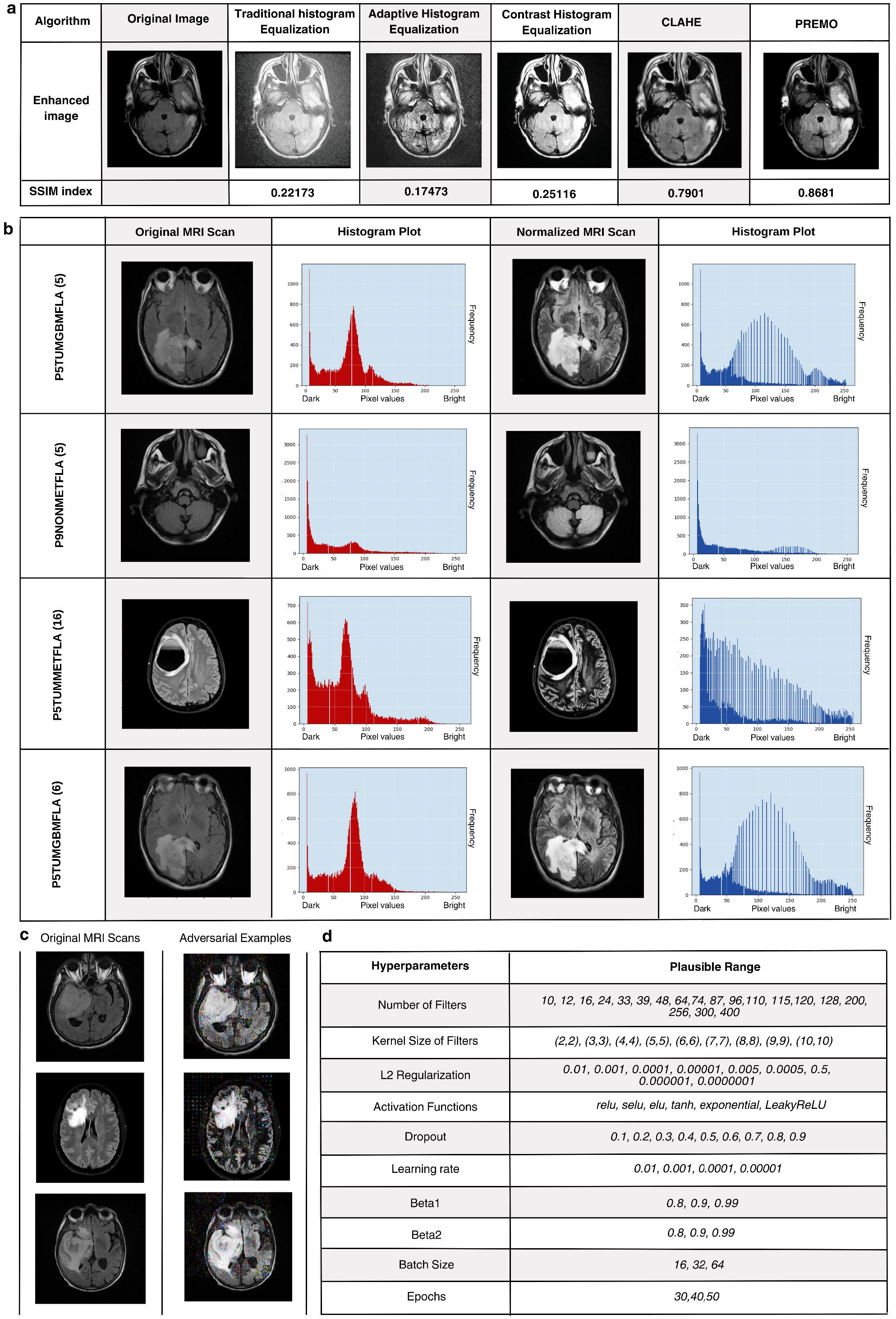
Contrast Enhancement and Performance Optimization of SIENNA. a. SSIM index of PREMO compared with pre-existing traditional histogram-based equalization algorithms show an approximate increase of 78 percent, indicating higher retention of scan complexities. SSIM index takes into consideration structural and textural information to quantify the similarity between unprocessed scans and equalized scans. b. PREMO contrasts equalized MRI scan examples, with plots showing redistribution of otherwise imbalanced intensities. c. Original scans alongside adversarial counterparts produced through normalized gradient perturbations scaled by a factor of *ε*. The magnitude of the perturbation was chosen to be 0.1, convolved with the gradient of the categorical cross-entropy loss of input image to produce each adversarial example. d. Parameter search spaces are pruned using HYPERAS.

To avoid data overfitting that can occur with small datasets and to enhance SIENNA’s ability for class discrimination, to differentiate between relevant and irrelevant features, we applied adversarial training samples [57,58] generated using the Fast Gradient Sign Method (FGSM) algorithm (see **Methods**). In short, the process involves calculating the gradients of the loss between the model’s prediction and the true value for a test case. Perturbations are then applied to the clean samples in the direction of the gradient, aiming to increase the loss, generating adversarial examples as shown (**Fig. 2c)**. The adversarial training samples are crafted in a manner to exploit the model’s vulnerabilities and decision boundaries while minimizing noticeable visual alterations that can add excessive noise and instead undermine the model’s ability to identify meaningful patterns and attributes. SIENNA’s optimal performance strikes the necessary balance between the quantity and characteristics of noise introduced into the training dataset and hyperparameter tuning during analysis (**Fig.2d**).

### SIENNA is a Generalizable Framework for Robust Multi-Classification of MRI Data

SIENNA’s co-development with minimally processed MRI DICOM data strengthens its generalizability and diagnostic capabilities relevant to clinical implementation. We analyzed de-identified patient data, 8 MET and 9 GBM-typed individuals, including similar numbers of male and female patients, **(Fig.3a)** showcasing a distribution of patient demographics, detailing aspects for age, sex, weight, and MRI machine manufacturer and magnetic field (1.5T - 3.0T) utilized. Patient ethnicity in the Albany Medical College data sampling was primarily Caucasian with one Black patient. The channelization of MRI patient data into the SIENNA workflow is summarized in **Fig.3b** and encompasses data histogram equalization by PREMO, the contribution of SMOTE and adversarial samples to training, hyperparameter tuning of training/validation/testing to achieve optimized performance metrics and final evaluation metrics. SIENNA’s objective and numerical assessments were cross-validated using a 100 repeated random sub-sampling method in non-interdependent multi-subtask classification analysis (**Fig.3c**). SIENNA’s results on clinical DICOM MRI data across 3 tasks have average accuracies of 92 percent (Non-Tumor, SD=5.5 percent), 91 percent (GBM, SD = 3.2 percent), and 93 percent (MET, SD = 2.6 percent). SIENNA’s non-interdependent subtasks allow additional identification of True Positive (TP) and True Negative (TN) and misclassifications of False Positive (FP) and False Negative (FN) for minimization. Metrics such as accuracy, F1 score, and Area Under the Receiver Operating Characteristic Curve (AUROC) [70] are used to further track performance improvement. The use of diverse quantitative metrics in the confusion matrix that align with relevant clinical performance metrics gives the model user greater information for diagnostic decisions and potential health outcomes.

SIENNA was compared to SOTA AI tumor detection models generated on public BraTS to see if they are generalizable to our clinical dataset. BraTS MRI high-quality dataset is prominently used in developing MRI AI platforms but has applied extensive pre-processing techniques like skull stripping, bias field correction, and noise reduction. The tumor dataset consists of NIfTI files divided into four different MRI modalities, each containing 155 sliced images of dimension 240×240×3. To best compare SIENNA versus other MRI SOTA AI models, either in binary or multi-classification, we first retrained all models to the new data and then ran our analysis. This allows us to evaluate whether the architecture itself is underperforming. SIENNA’s ability to determine the absence or presence of tumors versus other MRI AI platforms was evaluated on a subset of the BraTS2020 dataset consisting of skull-stripped data from 20 patients. SIENNA, SCENIC, and NeuroXAI notably excelled in accuracy and AUROC, with values reaching up to 0.99, significantly surpassing CNN-SVM’s [44] peak accuracy of 0.78. (**Fig.3d**). Having compared the performance of models on processed data, we now evaluated models on our clinical data (GBM-MET). A contrast in performance emerged as SIENNA retained a high degree of accuracy, only slightly reduced from its performance on the processed data. On the other hand, SCENIC experienced a notable decline, and Xception’s metrics plummeted dramatically, suggesting potential overfitting to their original training datasets. CNN-SVM, although not the frontrunner on processed data, displayed a consistent performance across both datasets (as summarized in **Fig.3e**). To further challenge the capabilities of SOTA models, we evaluated their performance in a more clinical tumor typing task, distinguishing between GBM, MET, and non-tumor classifications (as shown in **Fig.3f**). In evaluations, SIENNA, calibrated for clinical datasets registered an accuracy of 0.91. Conversely, Xception, originally trained on the ImageNet corpus, demonstrated diminished proficiency, and CNN-SVM, notwithstanding its adeptness with the BraTS dataset, yielded a modest accuracy of 0.50 within the clinical paradigm, all highlighting SIENNA’s high accuracy of up to 91 percent and demonstrated the inability of other SOTA models to generalize to this more complex clinical dataset. As evident in the histogram comparisons, SIENNA outperforms other compared AI models and demonstrates the greatest stability, that is its ability to evaluate across different datasets and processing, reflected in a low SD.

**Fig. 3.**
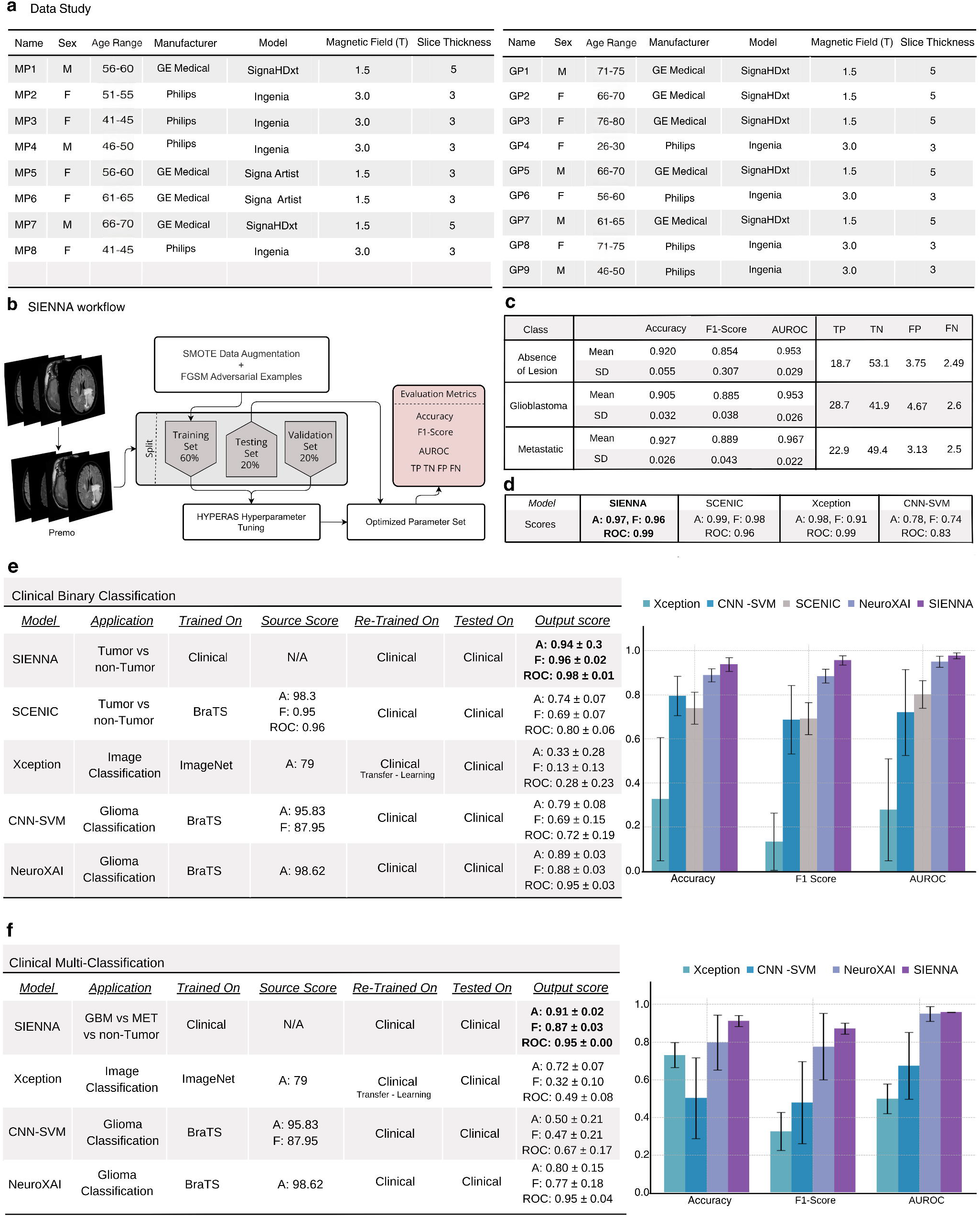
Clinical Data Study and Comparative Analysis of SIENNA with State of the Art. SIENNA outperforms state-of-the-art with high-accuracy multi-classification tumor analysis on multiple dataset types. a. Summary of clinical data used for analysis of SIENNA. All patients are Caucasian, age categorized into non-overlapping five year brackets, and weight is in kilograms, 1.5 or 3.0 Tesla acquired on clinical General Electric and Philip’s MRI machines. b. The workflow of SIENNA integrates minimal data pre-processing, histogram equalization, and adversarial training in a hyper-parameter-tuned network to generate a range of useful performance metrics. c. SIENNA’s ability to detect GBM and MET in our minimally processed clinical dataset with an accuracy of 91 percent and 93 percent cross-validated across 100-fold. d. Performance comparison using 5-fold cross-validation for tumor detection of the BraTS dataset shows that SIENNA performs comparably with SCENIC and Xception and outperforms CNN-SVM (N=5). e. Performance comparison using 5-fold cross-validation for clinical binary classification reveals SIENNA’s strong accuracy, while SCENIC’s performance declines and Xception overfits to data. NeuroXAI performs decently but struggles with generalization. f. Performance Comparison of multi-classification using 5-fold crossvalidation: SIENNA achieved 91 percent accuracy in clinical tumor typing, while Xception shows reduced proficiency, and NeuroXAI demonstrates decent performance with an unstable accuracy of 80 percent (N=5).

### Generalizability of SIENNA for New Patients and Interpretability of Predictions

SIENNA’s development on a small DICOM dataset increases the challenges with complex physiology. This includes features such as white matter lesions, faint presence of tumors, and high presence of non-brain tissue, which are more likely to be misdiagnosed in small datasets. To better evaluate SIENNA data handling, we identified the most frequently occurring MR images across the 100 runs for TP, TN, FP, and FN outcomes (**Fig. 4a)**. We next performed a patient-personalized assessment of the model’s performance on individual cases (**Fig. 4b**) typical in clinical practice. A detailed breakdown of the model’s performance on a patient-specific level to correctly classify GBM and MET cases is provided in the evaluation metrics. In **Fig. 4b** (left), the distribution of TP, TN, FP, and FN metrics specific to patients within each bar is shown. In some patients when slices with tumors represent the bulk of data, no TN will be present, and if no FP or FN are detected then only TP is indicated. We further output SIENNA’s diagnostics on the complete set of Z-slices within a patient data profile (**Fig.4c**). This highlights more challenging regions for SIENNA diagnostics. The analysis of patient MRI tumor data by non-interdependent analysis of 2D axial slices from intermixed Z-sections provides the greatest opportunity for robust analysis.

**Fig. 4.**
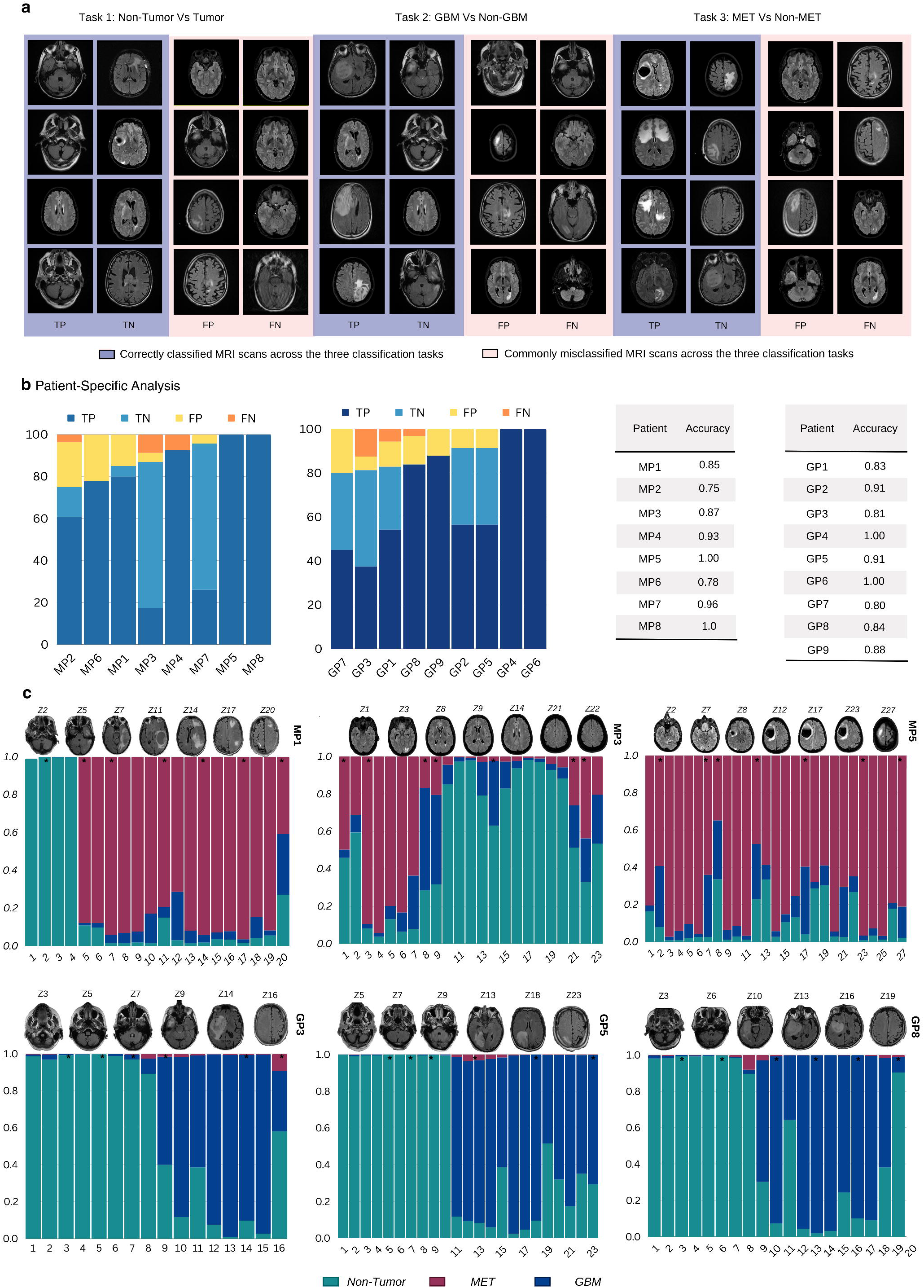
Personalized Evaluation of SIENNA’s Performance True to Clinical Setting. a. Analysis reveals that a limited set of false positive (FP) and false negative (FN) images with unique pathology characteristics and overbearing traces of the skull remain challenging for diagnostics. Extended training of SIENNA to larger datasets is expected to reduce these occurrences. b. Performance metrics are evaluated across patient-specific cases and organized based on accuracy. Stacked bars identify the proportion of accurate and misdiagnosed classifications, represented as TP (successful identification of positive class), TN (successful identification of negative class), FP (incorrect positive identification), and FN (incorrect negative identification). Patient-wise accuracy for GBM lies between 0.8-1.0 percent and for MET between 0.75-1.0 percent, where the accuracy is 1.0 when SIENNA detects no FP or FN. c. Confidence percentages mapped across all scans of a patient Z-stack with selected images shown above for three each representative MET (MP1, MP3, MP5) and GBM (GP3, GP5 and GP8) patients.

To understand class prediction by SIENNA, for tumor pathology features in GBM and MET diagnostics, we used Grad-CAM [71] analysis (**Fig. 5a**). This allows us to visualize gradients of the target class output concerning the feature maps of the last convolutional layer. Discriminative features were interpretably visualized by generating a heat map that identifies regions where the model may be over-relying or under-relying for the decision-making process for GBM (**Fig. 5b**) and MET (**Fig. 5c**) tumors. The presence of certain factors, pattern of spread, multiplicity, distinct signal characteristics, tumor size, and concentration in a particular area are indicative of a positive correlation with the occurrence or prediction of pathologies. In **Fig. 5b**, heat maps of GBM slices exhibit concentrated regions of interest. Misclassified GBM slices either fail to detect these pathologies or, when they do, often misclassify them as metastatic due to the presence of skull cavities or white matter, which may resemble tumors. Conversely, in **Fig. 5c**, MET Grad-CAMs tend to display a more spatial representation, accounting for tumor multiplicity in the decision-making process. Consistent with these observations, Grad-CAM analysis of MET slices that are incorrectly classified as GBM, show patterns that are tightly focused around specific brain instances, and the opposite also holds for GBMs misclassified as MET. Such analysis allows continued optimization of SIENNA through adversarial training and on expanded datasets.

**Fig. 5.**
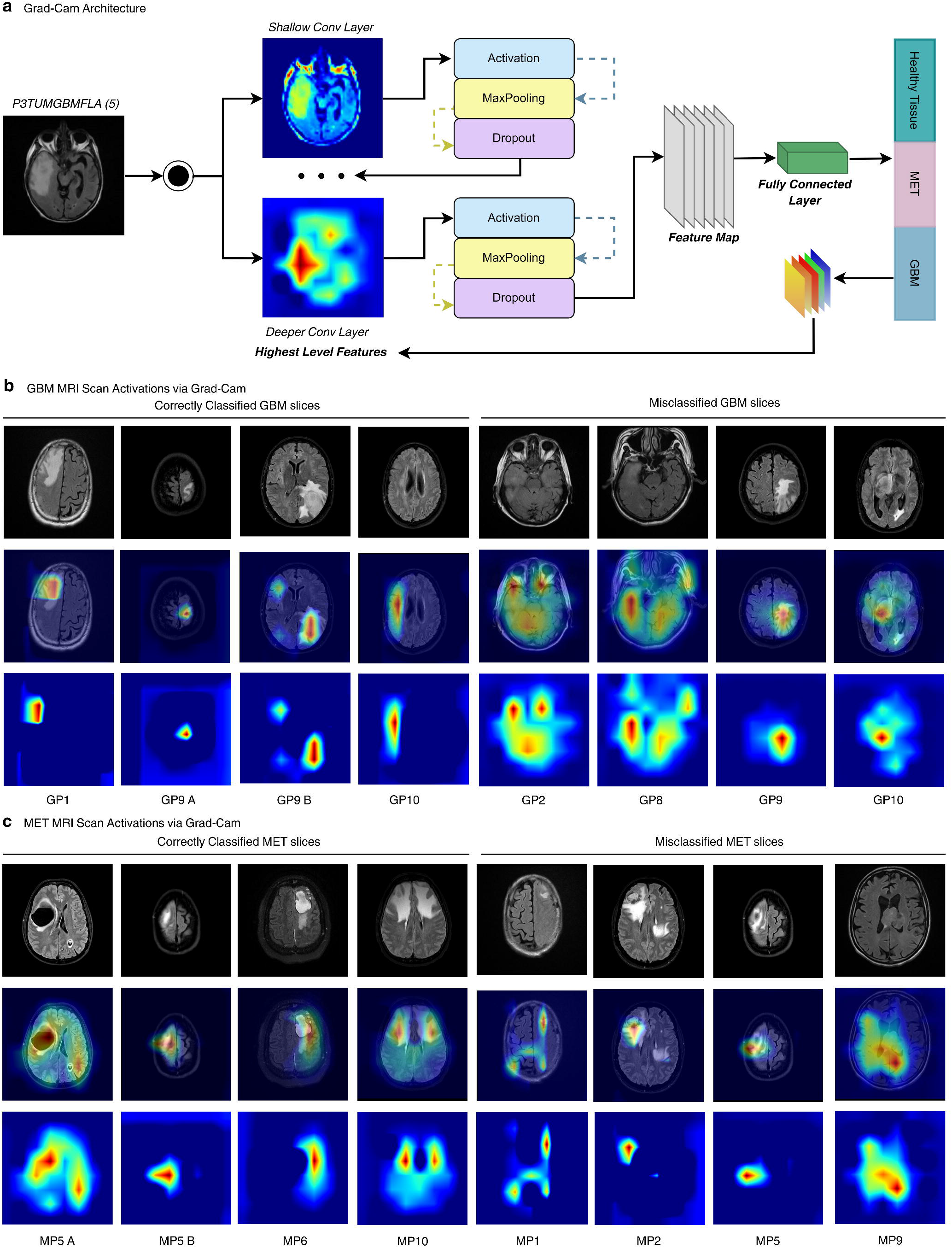
SIENNA Captures Discriminative Tumor Classification Features to Expedite Clinical Workflow. a. Grad-CAM architecture for explainability. Grad-CAM analysis reveals feature maps of the last convolutional layer to identify discriminative features. In terms of FP or FN, these regions inform on where the model may over-rely in decision output. Early decision capabilities for GBM and MET classification can alert pathologists to scan for primary cancers. Normally this action is delayed due to poor or unreliable MRI data and the need for additional diagnostics. b. Heat map analysis diagrams for GBM. c. Heat map analysis diagrams for MET.

SIENNA was next examined in regard to the number of trainable parameters versus various other MRI-based ML models developed over the past decade that reveal significant differences in model sizes (**Fig. 6a**; see also **Table S1**). We observe that SCENIC developed by our group in 2023 and SIENNA in 2024 in this study have significantly lower model size compared to other contemporary models. We next examined inductive classification performance of various models (**Fig. 6b)** and show that SIENNA achieves the highest performance while having the least number of parameters, i.e., being the smallest model. Hence, we establish the AGI hypothesis of small models achieving better generalizability [16] in image classification with a focus here on MRI-based tumor diagnostics.

**Fig. 6.**
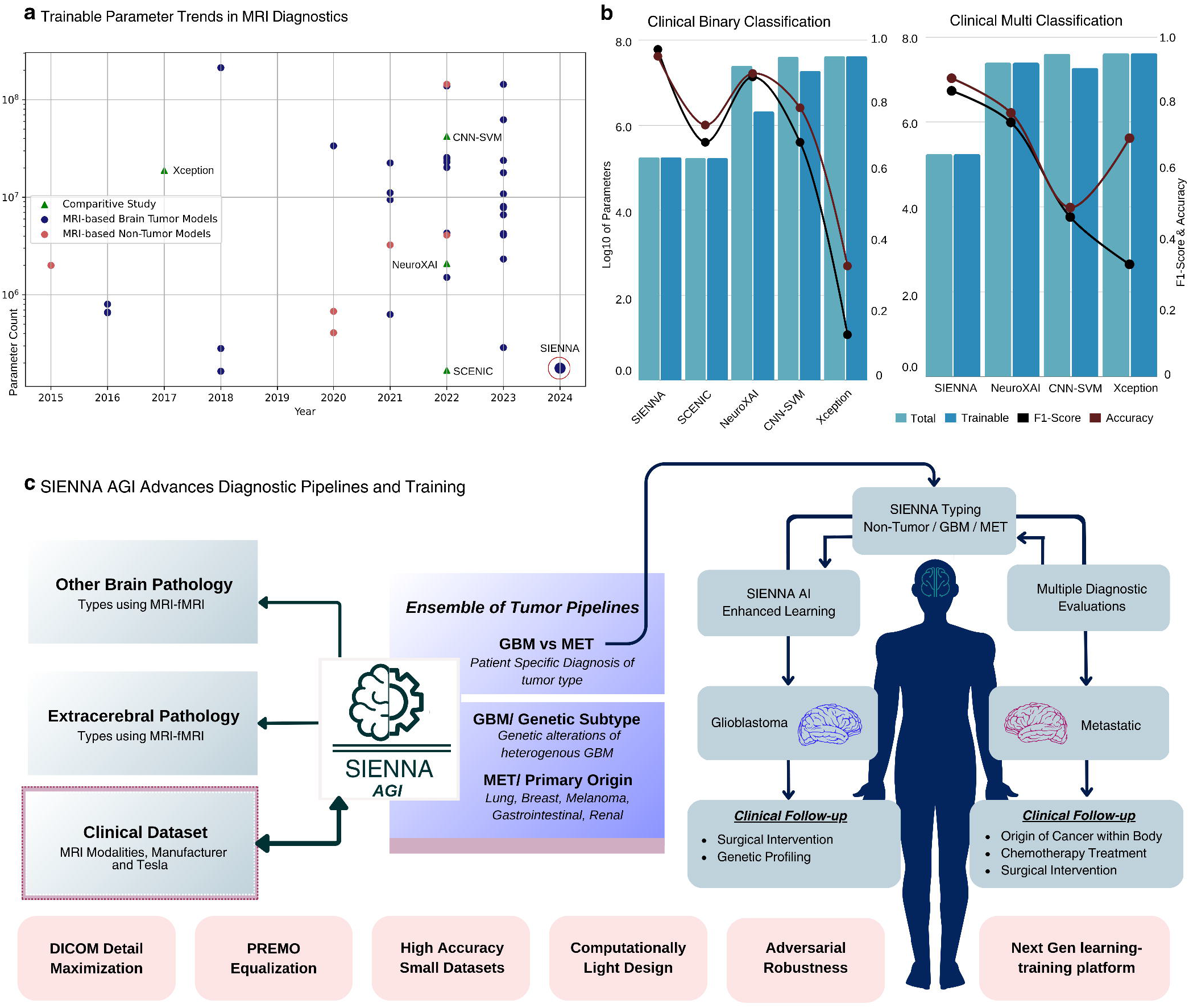
SIENNA AGI Integrates Data Handling and Multiclass Diagnostics. a. Trends in learnable parameter count from 2015 to 2024, across brain tumor and non-tumor related medical imaging applications b. Plot visualizes correlation between parameter count and accuracies of state-of-the-art models used for our comparative analysis. Line chart overlay visualizes F1 score and accuracy metrics. SIENNA demonstrates superior efficiency with fewer parameters. Conversely, Xception and CNN-SVM, despite their higher parameter counts, exhibit inferior performance compared to SIENNA. c. Overview of the current and expandable potential of SIENNA in diagnostic pipelines. SIENNA is a high-accuracy companion diagnostic for tumor multiclass classification. SIENNA’s ML architecture and integrated optimized data handling are optimal for the analysis of clinical DICOM MRI data and adaptability to different datasets and diagnostic pipelines. SIENNA also offers AI enhanced learning and training for clinical practitioners.

## DISCUSSION

AI is emerging as a companion diagnostic for clinical medical imaging, but challenges remain to mitigate shortcut learning, improve generalizability to new patients, enhance explainability, and evaluate challenging pathologies. Here we focus on addressing these challenges in the development of SIENNA, with particular attention to clinical MRI data handling, designing inductive TTV (train-test-validation) for improving generalizability, and reducing errors that are evident in brain tumor pathology diagnostics [9, 24, 25]. MRI ability to capture increasingly high-resolution physiological detail and incorporate continuing technological advancements make it indispensable to patient diagnostics and long-term care. We demonstrate via SOTA comparisons that SIENNA’s ML tumor diagnostics platform can generalize to new patients while trained on limited data. SIENNA output includes advances in multiple areas, summarized in **Fig. 6c** that are 1) improved clinical DICOM data handling that captures detailed brain neurophysiology features for ML training and validation, 2) capability to work with small clinical datasets, 3) generalizability to handle complex clinical data, over processed data, and new patients, 4) non-interdependent multi-classification of tumor type pathophysiology and tracking of FP, FN, TP, TN outcomes, 5) a computationally light, low memory consumption, and portable design to benefit clinical integration and team communication, 6) a buildable AGI framework for brain tumor diagnostics as well as an ensemble of future diagnostic pipelines, and 7) AI-enhanced learning and training for clinical practitioners.

SIENNA is novel in its generalizability to analyze data via a neural architecture that is integrated with minimal DICOM data preprocessing. This includes development of a histogram equalization algorithm, PREMO, optimized to retain feature details while minimizing noise. Attention to data handling in our design was critical to achieve a high inductive accuracy (91-93%) for unseen patients in multiclass typing of tumor presence, GBM, and MET neuropathologies in a small clinical dataset. By comparative studies we demonstrate that this approach uniquely allows SIENNA to be transferable, retaining high accuracy when tested with an alternate highly processed BraTS dataset. In contrast, the state-of-the-art ML models trained, and validated on highly processed MRI tumor data were not generalizable to our clinical dataset, resulting in significantly reduced accuracy, F1 and AUROC values. SIENNA also demonstrates greater statistical stability in decisions with extremely low standard deviations. Development of SIENNA’s generalizable capabilities is a first step in the planned future expansion of SIENNA to subtype tumors, such as GBMs with genetic signatures in a multimodal setting and MET analysis that informs on primary tumor source (**Fig. 6c**). By GradCam analysis (**Fig. 5**) SIENNA recognizes different features in MRI tumor GBM data versus METs as expected for detail oriented multi-classification. While NeuroXAI [51] uses visual attention maps for explaining MRI analysis of brain tumors, SIENNA can achieve similar explainability by identifying similar locations on the images via GradCam. We anticipate that increased training of SIENNA, particularly with METs of more complex physiology, will generate even higher accuracy for tumor multi-classification. To generate data outputs useful to neurosurgeons and radiologists we provide information on TP, TN and FP, FN outcomes per patient diagnosis. Prediction outcomes are performed per slice for Z-stack axial scans, that provide the clinical expert the ability to understand diagnostic decisions made by SIENNA. Our choice to work with 2D axial images versus generated 3D images enables SIENNA broader use, such as the small clinical dataset used here or integrated diagnostics during real time MRI data capture. Furthermore, the use of 2D axial images helps in reducing the model size and eases the deployment of the model in the clinical setting.

Our training and analysis with SIENNA used intermixed 1.5 and 3.0 T data from General Electric and Philips MRI machines respectively. Significantly this indicates that an AGI architecture enhanced by retaining dataset features that are critical to object representations need not be specific to proprietary algorithms that define raw MRI data. SIENNA architecture is focused on building algorithms that better understand the visual task differentiations needed in complex clinical data. SIENNA tumor multi-classification in small datasets does reveal challenges surrounding distinct pathology features such as white matter lesions, faint presence of tumor, high presence of non-brain tissue, and low resolution. These were minimized in our design and are expected to be further reduced with larger training sets to account for the patient-wise data variability. SIENNA’s non-interdependent multi-classification supports expanded subtyping classification to bring in genetic information on tumor types, such as glioblastomas, or potentially training SIENNA to recognize features that are indicative of metastasized tumor primary site. For the latter, MET studies indicate such physiologic correlations appear to exist [10,19-22]. Changes to magnetic power alone either to vastly increase feature detail as in 7.0 T data or use of lower Tesla MRI systems for more frequent monitoring will also increase demand for AGI companion diagnostics. Such data will be inherently noisier in detail or lack thereof emphasizing the continued demands on data handling. SIENNA’s ability to work with small clinical MRI DICOM datasets and data handling methodologies highlight an AI architecture poised to expand further in medical imaging diagnostic applications.

## METHODS

### Clinical Dataset Description

The de-identified clinical data used in this study for training, validation, and testing of SIENNA were obtained as part of an IRB-approved collaboration with Dr. Pilitsis (Albany Medical College IRB 6127) and includes 17 patients (for complete dataset description see Fig 3a). The MRI files encompass Tesla magnetic field strengths of 1.5T and 3T, generated by General Electric SIGNAHDxT 1.5T, GE SIGNA Artist 1.5T, and Philips Ingenia 3T MRI Machines. Thus, our cross-validation assesses SIENNA’s generalizability across a patient cohort of males/females spanning middle to older adulthood for GBM and MET, while encompassing different magnetic strengths and MRI machine manufacturers. All radiographs are axial plane slices and were provided in the Digital Imaging and Communications in Medicine, DICOM, format. The patient files were expert-typed as MET or GBM classes and included multiple 2D image scans (slices) within a 3D Z-plane series. To simplify metadata complexities and reduce image storage demands, we converted individual DICOM files to the Portable Network Graphics (.png) format and the DICOM metadata was re-annotated to adhere to a uniform labeling format for all 17 patients. The format includes information such as patient number, tumor presence (TUM for tumor and NON for non-tumor), tumor type (MET for metastasized and GBM for Glioma), modality type (FLA for FLAIR), and the slice number [P8TUMMETFLA(3)]. These identifiers are utilized post-analysis to re-align data outcomes from individual 2D images to the original patient files to benefit patient-specific analysis.

To reduce noise in a small dataset, we excluded from training and analysis the most outlying scans in a 3D axial patient Z-stack. This includes the uppermost scans which predominantly contain a portion of the cranial structure, and the lowermost scans which include the neck region. Such images were outside of the tumor identification regions. This was sufficient to reduce spatial intensity differences between skull and/or brain tissues in these regions that can mislead the model into learning incorrect patterns that can result in overfitting irrelevant features [72]. For example, the distinctive appearance of a fraction of the skull that has no or minimal brain tissue in one axial slice versus a midbrain axial slice can impede generalizability. This method is preferred versus over-processing methods such as skull-stripping of all data seen in public datasets. A total of 386 scans from 17 patients were sampled for SIENNA analysis including 153 GBM, 125 MET, and 108 non-pathological (non-tumor) slices. Slices of diverse dimensions (ranging from 356 x 356 to 528 x 528) were standardized by resizing to 240 x 240 pixels before being used as input for SIENNA using the OpenCV library [73]. The resizing ensures uniformity in size across all scans, which is essential for batch processing and memory management considerations. Although it may introduce aspect ratio distortion and smoothing, we did not observe a substantial reduction in downstream performance due to resizing.

### Model Training, Validation, and Testing

Evaluation of training performance and generalizability are both required in assessing machine learning (ML) models [74, 75] to ensure that a high training performance does not just reflect a model’s ability to memorize specific training data that may limit its ability to generalize to unseen data. Our validation set during SIENNA training measures the model’s accuracy on a dataset different from the training data and includes an inductive validation set that helps to tune the model parameters and hyperparameters for unseen data, avoiding overfitting [76]. We randomly split the clinical data into training (60 percent), validation (20 percent), and test (20 percent) datasets and shuffled the MRI scan slices between these image datasets to minimize the chance of SIENNA unintentionally capturing a subset of patient-specific characteristics or becoming dependent on select images or sequential patterns linked to the patients, thus aiding generalizability.

For SIENNA training, the original MRI dataset consists of 233 slices, with an average distribution of class labels across 100 random iterative splits 71 non-tumor, 88 GBM, and 74 MET slices. To mitigate the class imbalance, we employed SMOTE augmentation [66] to resample the under-represented MET and non-tumor classes, applied to 40% of minority class samples, and generated new synthetic data. SMOTE rebalances classes by generating new data examples from original data by interpolation of feature space unique to minority classes. In a k-nearest neighbor (KNN), ML approach (KNN) Euclidean Distance is used to represent regional features in n-dimensional space, such that the value of a feature is dependent on KNN with k=8 nearest neighbors in our process. For each pair between the original point and its neighbor, we interpolate the intensities to create synthetic images. These images are inserted randomly within the minority class data to restore class balance for SIENNA during training and testing, followed by performance validation across 100 non-interdependent data splits (Fig. 3d).

To enhance SIENNA’s performance, we implemented a targeted augmentation feedback loop, which includes the iterative identification of frequently misclassified MRI scans and their subsequent incorporation into the training data. An FP-FN occurrence count is kept for each scan ID, and once it surpasses a predefined threshold of 15, we permanently incorporate these scans into the training process. Targeted augmentation ensures that SIENNA, observing 4.2 percent of images drawn from a distribution different from the training data, can generalize well to unseen and unique pathology test cases. Adapting the model’s learning process to accommodate these distinctive data points enhances SIENNA’s robustness, transferability, and generalizability to new patients.

### Intensity normalization across patient data

Comparability of data is critical and to what degree MRI machine source that includes proprietary algorithms to convert raw data of MR into images, acquisition expertise and parameters (slice thickness, echo time), and tesla intensity variations (1.5-3.0) impact comparability is not fully known [77]. Intrinsic variations in pixel intensities for the same tumor pathologies and within the same modality across patients are common. This variation can create challenges for deep learning models in generalizability for pathology companion diagnostics. To address these challenges and determine the most robust method for data harmonization in MRI image datasets for SIENNA analysis we incorporated histogram equalization [56, 67] which is known to benefit model accuracy and generalizability [78, 79]. In this study, we applied and validated several histogram equalization techniques before developing our own improved method, referred to as PREMO (Fig. 2a). This includes Adaptive Histogram Equalization (AHE) [56], Contrast Histogram Equalization (CHE) [68], and Contrast Limited Adaptive Histogram Equalization (CLAHE) [68] which were tested and compared using the Structural Similarity Index (SSIM). In use with SIENNA, these approaches exhibit limitations that hinder their effectiveness in improving image quality and accurately detecting and characterizing tumors. A key limitation of traditional histogram equalization methods is their insufficient preservation of brain structure [80]. These methods tend to enhance the entire image uniformly, including non-brain regions, which compromises the visibility of critical brain structures (as seen in Fig 2a). Further, the existing algorithms demonstrate ineffective handling of variable intensity distributions. AHE and CHE algorithms are designed to enhance local contrast based on local intensity distributions and do not adequately adapt to the variable intensity distributions present in brain MRI scans. These limitations lead to inconsistent and suboptimal enhancements, further impacting the quality of the images. Moreover, the existing algorithms lacked sufficient control over enhancement parameters. Although CLAHE, a variation of AHE [56], does address some of the limitations of traditional methods by limiting contrast enhancement, it still lacks the flexibility to selectively apply enhancements based on specific regions of interest. Building upon these existing algorithms and inspired by CLAHE improvements, we introduce PREMO, a method tailored for redistributing pixels across intensity levels, improving equalization through masking, and optimizing brightness and contrast via Gamma fine-tuning in clinical MRI datasets. Specifically, PREMO is designed to efficiently isolate the region of interest from the background, applying adaptive contrast enhancement between tumor pathology and tissue. This process enables control over brightness and intensity redistribution, effectively scaling the image to an intensity range of 0-255. We consider an MR image, represented as M(p, k), where p would typically be represented as a tuple (*x*_*p*_, *y*_*p*_), p belongs to *R*^*2*^, representing the 2D spatial coordinate of the image, denotes the pixel position coordinates and k represents the color channel. Since MR images are typically grayscale, Otsu’s thresholding [81] technique is applied to convert the grayscale input image into a binary image. Otsu’s method determines an optimal threshold value by maximizing the between-class variance. That value is used for binary thresholding operation, which can be expressed mathematically as follows:

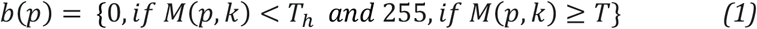

where b(p) denotes the binary image pixel value at position coordinates p and *T*_*h*_ represents the threshold value determined by Otsu’s method, which selects a threshold in such a way that it minimizes intraclass variance between the foreground and background of the scan. Following thresholding, morphological opening is performed on b(p) using a 5x5 kernel, which suits a balance between noise removal and preserving brain area to further refine the image and eliminate noise, which is used to create a mask of the scan to distinguish the background and brain area. The morphological opening operation can be mathematically represented as:

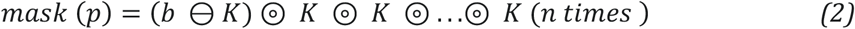

Here, mask(p) represents the resulting mask image, b denotes the binary image obtained through thresholding, K represents the 5x5 kernel, and n denotes the number of iterations (in this case, 3 iterations) for the morphological opening operation. The morphological opening operation involves erosion (⊖) followed by dilation (⊚), which removes small noise elements and smoothens the regions of interest. Erosion erodes the boundaries of the foreground regions in the binary image, while dilation expands the boundaries back using the same kernel, eliminating small gaps or holes.

Variations in pixel intensity range can occur due to differences in MRI machine software or other parameters [82], where the intensity range is represented as [*G*_0_, (*G*_L_ − 1)], where *G*_0_ is the minimum intensity value and *G*_L_ − 1 is the maximum intensity value. The probability density function (PDF) of intensity i (Eqn. 3) provides details about the distribution and occurrence patterns of various pixel intensity levels within and across the MRI images. The cumulative probability function (CDF) generates a comprehensive measure of the accumulated probabilities associated with observing different intensity levels across the entire range of the MR image (Eqn. 4) [83]. The traditional histogram equalization transformation function T (Eqn. 5) maps the CDF of each pixel of M (i, k) to a uniform distribution to produce the equalized scan *Eq*_*img*_(Eqn. 6).

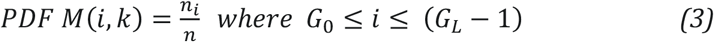

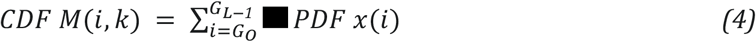

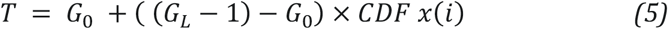

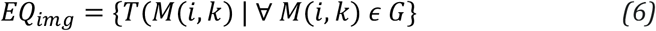

where,

*n*_*i*_ represents the total number of pixels with intensity value i in M(i, k) n is the total number of pixels

G represents a set of possible values for M(i, k), which are the pixel intensity values in the image

Subsequently, a contrast stretching operation is applied to normalize the pixel values within the range of 0 to 255, thereby enhancing the image contrast [84]. This operation aims to optimize the visual appearance of the image by adjusting the pixel intensities. The contrast stretching operation is expressed by the following equation:

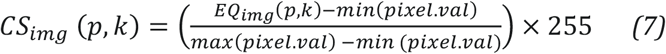

where, min(pixel. val) and max(pixel. val) represent the darkest and brightest pixel intensity values, respectively, in the equalized image after applying the histogram equalization process.

Here, *CS*_*img*_(*p, k*) in Eqn. 7 denotes the contrast stretched pixel value in the output image. Furthermore, to improve the overall brightness of the image and achieve fine control over its visual appearance, we apply a Gamma correction [74]. Gamma correction in PREMO involves raising each pixel value to the power of the specified γ value. Gamma correction is expressed by the equation:

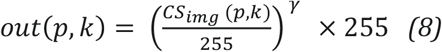

In Eqn. 8, *out*(*p, k*)) represents the pixel value in the output image and γ signifies the value used for the correction, set γ to 3. By incorporating the contrast stretching and Gamma correction technique used in PREMO, we can enhance the contrast and adjust the overall brightness of the MRI image.

To confirm the retained quality of processed images after histogram equalization, we have utilized the Structural Similarity Index (SSIM) [57] to evaluate the structural similarity between the original scans (M(p,k)) and the corresponding enhanced scans (out(i, j)). SSIM Index is computed as.

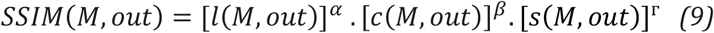

where l(M, out), c(M, out), and s(M, out) represent the luminance, contrast, and structure components, respectively, and α, β, and Г are weighting parameters that determine the relative importance of each component. SSIM index of PREMO enhanced scan is found to be 0.86 (**Fig. 2a**). SSIM values range from -1 to 1, with a value of 1 indicating perfect structural similarity between two images. An SSIM of 0.86 suggests a substantial degree of structural congruence between the processed and reference images. However, it is imperative to note that this metric neither conveys a linear percentage similarity nor provides insight into the image’s smoothness or sharpness. Instead, it focuses on structural information preservation.

### SIENNA Architecture

SIENNA is a portable and computationally efficient platform capable of 1) processing clinical data on par with high-accuracy and low FP/FN standards necessary in clinical companion diagnostics, and 2) accurately detecting tumor presence and categorizing the pathology as either GBM or MET. The architecture of SIENNA is outlined in Fig. 1e and the workflow is detailed in Fig. 3b. Data preprocessing has been described and includes exclusion of outlying slices, PREMO histogram equalization, and SMOTE augmentation to restore class balance. SIENNA employs a multilayer-linear CNN architecture for feature extraction, constructed using the Sequential API in TensorFlow/Keras. This architecture comprises 12 feature extraction layers, encompassing convolutional layers, activation layers, max-pooling layers, and dropout layers. Additionally, it comprises four classification layers, including flattened, dense, dropout, and output layers. The SoftMax activation function is applied to produce class probabilities, estimating the class-specific probability for each input MRI scan slice. These layers and feature parameters are tuned using hyperas [60] (Fig. 2d) and demonstrate an efficient exploration-exploitation tradeoff [75].

Given SIENNA’s multi-class functional nature, categorical cross-entropy is chosen as the primary loss function for error estimation. Moreover, we have incorporated false positive (FP) and false negative (FN) metrics as auxiliary loss functions during the tuning process. This is especially pertinent in the realm of medical diagnostics, where the consequences of FP and FN outcomes can be of substantial risk to the patient’s health [76]. The composite training loss function (*C*_*train*_) is formulated as follows for an n-class classification task:

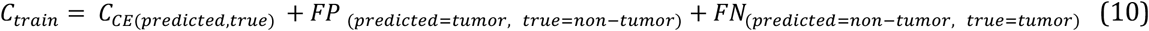

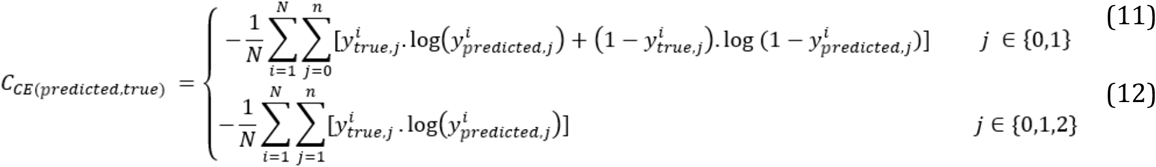

where N is the number of samples in the MRI dataset, n is the number of classes, 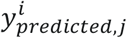 is the true label of the class *j* of the *i*^*th*^sample.

Eqn. (11) and Eqn. (12) represent the binary cross-entropy and categorical cross-entropy used for tumor detection (binary classification) and typing (multi-type classification), respectively.

We apply the Adam stochastic gradient descent [88] algorithm for iterative parameter optimization. This choice of the optimizer is made through an exhaustive exploration across various stochastic optimizers [88] such as stochastic gradient descent (SGD) [90], Root Mean Square Propagation (RMSprop) [91], Adaptive gradient descent (adagrad) [92]. We implement early stopping [93] for each training instance during hyperparameter tuning to terminate the training process if the validation performance remains unchanged or degrades in 15 consecutive epochs. The training process is conducted on a Microsoft Windows 11 workstation equipped with an Intel(R) Core (TM) i7-10750H six-core CPU, 16 GB of system RAM, and a single NVIDIA GTX 1650 GPU boasting 4 GB of GPU RAM.

### Mitigating Overfitting in Small Datasets Using Adversarial Examples

To enhance and optimize SIENNA generalizable diagnostics across diverse patients’ image datasets, we tackle the issue of overfitting [94] by introducing adversarial training [95]. This method introduces perturbations in the training data, which enables SIENNA to handle small variations in feature input and perform effectively with new MRI data. We use the variable *h*_θ_ to represent the neural network model, *x*_*adv*_ represent the generated adversarial example, and *y*_*true*_ represent the true label. The goal is to maximize the loss function via generated adversarial examples that improve SIENNA’s performance. Here we utilize the Fast Gradient Sign Method (FGSM). By calculating the loss gradient of the model, we produce perturbations in the direction of this gradient, which are intended to maximize the loss, and then apply these over MRI data samples. Randomly selected seed samples from a shuffled dataset are used for a forward pass to obtain the predicted labels. This can be represented as:

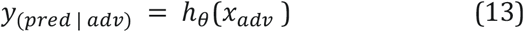

The loss function (Eqn. 13) which quantifies the discrepancy between the true labels and the predicted *y*_*pred* | *adv*_, varies based on the classification type, as calculated by (11) and (12).

Given this loss function (Eqn. 13), the gradient which pinpoints the direction where *x*_*adv*_ should be modified to maximize the discrepancy between the true label and *grandients* (Eqn. 14) can be represented as:

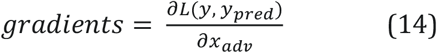

To ensure consistent perturbation magnitudes, we normalize these gradients using a small constant, (*ϵ*_1_), to prevent division by zero and is represented (Eqn. 15) as:

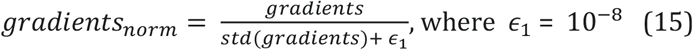

Following this, the adversarial example *x*_*adv*_ is updated by adding the element-wise product of the gradient gradients and the perturbation strength *ϵ*. This can be represented mathematically in Eqn. 16 as:

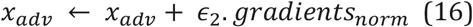

where *ϵ*_2_ represents the perturbation strength, with the value set to 0.1. This step allows us to manipulate the MR image data input in a targeted manner to produce perturbed examples that are designed to purposefully mislead the SIENNA’s predictions. By applying this perturbation, we not only explore SIENNA’s sensitivity to small changes in the MR image data but also focus on the key identifiable, discriminative features, ignoring irrelevant noise, and ensuring that SIENNA is robust against types of noise it might misclassify in the test of the MRI data set. Although this technique improves the generalization ability of the CNN model in SIENNA, a trade-off is mandatorily needed between the robustness and the accuracy, where an overly robust model may decrease accuracy on normal MR image data input [96].

## DATA AVAILABILITY

Access to clinical data is restricted to ensure patient privacy and confidentiality. The BraTS dataset is available at https://www.med.upenn.edu/cbica/brats2020/registration.html.

## CODE AVAILABILITY

Access to the complete source code for SIENNA is restricted due to existing patents. A Python script outlining essential pre-processing steps for MRI test slices (including PREMO, which is patented) and pre-trained weights of SIENNA is available at https://github.com/ITrakNeuro/SIENNA-Nature.git.

## Data Availability

All data produced in the present study are available upon reasonable request to the authors and the code is available on our GitHub repository.

https://github.com/ITrakNeuro/SIENNA-Nature.git

## ACKNOWLEDGEMENTS

JLP, AM, and AC conceived the project and experimental design and contributed to the generation and analysis of the SIENNA architecture, data analysis, and writing of the manuscript. JGP provided de-identified clinical MRI data and expertise. AC supervised data preprocessing and ML metrics. SSK and RSR generated the software, performed analysis, and developed figures with assistance from all co-authors. All authors contributed to the article and approved the submitted version. SIENNA analysis is supported by the State University of New York Research Foundation Technology Accelerator Fund (TAF) award.

## SUPPLEMENTAL TABLE S1

Supplemental Table. 1 Details of state-of-the-art Tumor Classification

## References

1. American Society of Clinical Oncology (ASCI) Brain Tumor Statistics (2023).

2. Ostrom QT, Cioffi G, Gittleman H, et al. CBTRUS Statistical Report: Primary Brain and Other Central Nervous System Tumors Diagnosed in the United States in 2012-2016. Neuro Oncol. 21, 1–100 (2019).

3. Brant-Zawadzki M. MR imaging of the brain. Radiology 166, 1–10 (1988).

4. Felix R, Schörner W, Laniado M, et al. Brain tumors: MR imaging with gadolinium-DTPA. Radiology 156(3), 681–688 (1985).

5. Edelman, RR. The history of MR imaging as seen through the pages of Radiology. Radiology 273, Suppl S161–S181 (2014).

6. Grand View Research. Magnetic Resonance Imaging (MRI) Market Size, Share & Trends Analysis (ID: 4076532) (2023). https://www.grandviewresearch.com/industry-analysis/magnetic-resonance-imaging-market

7. Miller, KD, Ostrom, QT, Kruchko, C, Patil, N, Tihan, T, Cioffi, G, Fuchs, HE, Waite, Ka Jernal, A. Siegel, RL, and Barnholtz-Sloa, JS Brain and other CNS tumor statistics. CA Cancer J Clin. 71, 381–406. (2021).

8. Saha A, Ghosh SK, Roy C, Choudhury KB, Chakrabarty B, Sarkar R. Demographic and clinical profile of patients with brain metastases: A retrospective study. Asian J Neurosurg, 8, 157–161 (2013).

9. Yan, P-F, Yan, L, Zhang, Z, Salim, A, Wang, L, Hu, T-T, and Zhao, H-Y. Accuracy of conventional MRI for preoperative diagnosis of intracranial tumors: a retrospective cohort study of 762 cases. Int J Surgery. 36. 109–117 (2016).

10. Schroeder T, et al. Mapping distribution of brain metastases: does the primary tumor matter? J Neurooncol. 147(1), 229–35 (2020).

11. Omuro A, DeAngelis DM. Glioblastoma and other malignant gliomas: A clinical review. JAMA. 310, 1842–1850 (2013).

12. Geirhos, R., Jacobsen, JH., Michaelis, C. et al. Shortcut learning in deep neural networks. Nat Mach Intell 2, 665–673 (2020). 10.1038/s42256-020-00257-z

13. Abdusalomov AB, Mukhiddinov M, Whangbo TK. Brain Tumor Detection Based on Deep Learning Approaches and Magnetic Resonance Imaging. Cancers (Basel). 2023 Aug 18;15(16):4172. doi: 10.3390/cancers15164172. PMID: 37627200; PMCID: PMC10453020.

14. Roelofs R, et al. A Meta-Analysis of Overfitting in Machine Learning. Advances in Neural Information Processing Systems 32 (NeurIPS 2019)

15. Brown, A., Tomasev, N., Freyberg, J. et al. Detecting shortcut learning for fair medical AI using shortcut testing. Nat Commun 14, 4314 (2023). 10.1038/s41467-023-39902-7

16. Sharma M et al., Smaller Models, Better Generalization. arXiv: 10.48550/arXiv.1908.11250

17. Xie L et al., Towards AGI in Computer Vision: Lessons Learned from GPT and Large Language Models, arXiv: https://arxiv.org/abs/2306.08641

18. Dolecek TA, Propp JM, Stroup NE, Kruchko C. CBTRUS statistical report: primary brain and central nervous system tumors diagnosed in the United States in 2005-2009. Neuro Oncol. 14, supp 5,v1–v49. (2012).

19. Delattre JY, Krol G, Thaler HT, Posner JB; Distribution of brain metastases. Arch Neurol 45(7).741–744 (1988).

20. Quattrocchi CC, Errante Y, Gaudino C, Mallio CA, Giona A, Santini D et al. Spatial brain distribution of intra-axial metastatic lesions in breast and lung cancer patients. J Neurooncol 110(1). 79–87 (2012).

21. Hengel K, Sidhu G, Choi J, Weedon J, Nwokedi E, Axiotis CA et al. Attributes of brain metastases from breast and lung cancer. Int J Clin Oncol 18(3). 396–401 (2013).

22. Takano K, Kinoshita M, Takagaki M, Sakai M, Tateishi S, Achiha T et al. Different spatial distributions of brain metastases from lung cancer by histological subtype and mutation status of epidermal growth factor receptor. Neuro Oncol 18(5). 716–724 (2016).

23. Tamimi AF, Juweid M. Epidemiology and Outcome of Glioblastoma. In: De Vleeschouwer S, editor. Glioblastoma [Internet]. Brisbane (AU): Codon Publications. (2017)

24. Morgenstern, JD, and Staudt, MD. Case report: Brain metastasis masquerading as glioblastoma multiforme and lymphoma. UWOMJ. 85, S1–S3 (2016).

25. Gaillard F, Rasuli B, Kang O, et al. Glioblastoma versus cerebral metastasis. Radiopaedia.org. doi: 10.53347/rID-43344 (2016).

26. Schiff D. Single Brain Metastasis. Curr Treat Options Neurol. 3, 89–99 (2001).

27. Blanchet L, Krooshof PW, Postma GJ et-al. Discrimination between metastasis and glioblastoma multiforme based on morphometric analysis of MR images. AJNR Am J Neuroradiol. 32, 67–73 (2011).

28. Cha S, Lupo JM, Chen MH et-al. Differentiation of glioblastoma multiforme and single brain metastasis by peak height and percentage of signal intensity recovery derived from dynamic susceptibility-weighted contrast-enhanced perfusion MR imaging. AJNR Am J Neuroradiol. 28, 1078–1084 (2007).

29. Essig M, Nguyen TB, Shiroishi MS et-al. Perfusion MRI: the five most frequently asked clinical questions. AJR Am J Roentgenol. 201, W495–510 (2013).

30. Fink KR, Fink JR. Imaging of brain metastases. Surg Neurol Int. 4, S209–S219 (2013).

31. Lapointe S, Perry A & Butowski NA. Primary brain tumors in adults. Lancet. 392. 432–436 (2018).

32. CADTH Report / Project in Briefs [Internet]. Ottawa (ON): Canadian Agency for Drugs and Technologies in Health; 2011-. Comparing 1.5 Tesla with 3.0 Tesla in Magnetic Resonance Imaging (2011).

33. Barisano G, Culo B, Shellock FG, Sepehrband F, Martin K, Stevens M, Wang DJ, Toga AW, Law M. 7-Tesla MRI of the brain in a research subject with bilateral, total knee replacement implants: Case report and proposed safety guidelines. Magn Reson Imaging (2019).

34. Harisinghani, MG, O’Shea, A, & Weissleder, R. Advances in clinical MRI technology. Science Transl Med. 11 (2019).

35. Deoni, S. C. L., Medeiros, P., Deoni, A. T., et al. Development of a mobile low-field MRI scanner. Nat. Methods 19, 569–582 (2022).

36. Oren, O, Gersh, B, Bhatt, DL. Artificial intelligence in medical imaging: switching from radiographic pathological data to clinically meaningful endpoints. Lancet. 2, e486–489 (2020).

37. Zhou, K, Greenspanm H, Davatzikos, C, Duncan, JS, Van Ginneken, B, Madabhushi, A, Prince, JL, Rueckert, D, and Summers, RM. A Review of Deep Learning in Medical Imaging: Imaging Traits, Technology Trends, Case Studies With Progress Highlights, and Future Promises. Proceedings of the IEEE. Institute of Electrical and Electronics Engineers (2021).

38. B. H. Menze, A. Jakab, S. Bauer, J. Kalpathy-Cramer, K. Farahani, J. Kirby, et al. The Multimodal Brain Tumor Image Segmentation Benchmark (BRATS). IEEE Transactions on Medical Imaging 34. 1993–2024 (2015).

39. S. Bakas, H. Akbari, et al. Advancing The Cancer Genome Atlas glioma MRI collections with expert segmentation labels and radiomic features. Nature Scientific Data (2017).

40. Smith, J., Johnson, A., Brown, R., & White, L.. Digital Imaging and Communications in Medicine (DICOM) Standard. Journal of Medical Imaging 20(6). 1243–1250 (2005).

41. Rorden C, Brett M. Stereotaxic display of brain lesions. Behavioral neurology. 191–200 (2000).

42. Tustison NJ, Avants BB, Cook PA, et al. N4ITK: improved N3 bias correction. IEEE Trans Med Imaging. 1310–1320 (2010).

43. Chakrabarty, S, Sotiras, A, Milchenko, MPamela LaMontagne, Michael Hileman and Daniel Marcus, D. MRI-based Identification and Classification of Major Intracranial Tumor Types Using a 3D Convolutional Neural Network: A Retrospective Multi-Institutional Analysis Radiology: Artificial Intelligence. 3 (2021).

44. Latif, G, Brahim, GB, Iskandar DNFA, Bashar, A, and Alghazo, J. Glioma tumors’ classification using deep-neural-network-based features with SVM classifier. Diagnostics. 12 (2022).

45. Hsu, W-W, Guo, J-M, Pei, L, Chiang, L-A, Li, Y-F, Hsiao, J-C, Colen, R, and Liu, P. A weakly supervised deep learning-based method for glioma subtype classification using WSI and mpMRIs. Sci Rep. 12, 6111 (2022) doi: 10.1038/s41598-022-09985-1

46. B. S. T. Naidu et al. SCENIC: An Area and Energy-Efficient CNN-based Hardware Accelerator for Discernable Classification of Brain Pathologies using MRI, 2022 35th International Conference on VLSI Design and 2022 21st International Conference on Embedded Systems (VLSID) (2022).

47. Chollet, F. Xception: Deep learning with depth wise separable convolutions. In Proceedings of the IEEE Conference on Computer Vision and Pattern Recognition (CVPR). 1800–1807 (2017).

48. Szegedy, C., Vanhoucke, V., Ioffe, S., Shlens, J., & Wojna, Z. Rethinking the inception architecture for computer vision. In Proceedings of the IEEE Conference on Computer Vision and Pattern Recognition (CVPR). 2818–2826 (2016).

49. He, K., Zhang, X., Ren, S., & Sun, J. Deep residual learning for image recognition. In Proceedings of the IEEE Conference on Computer Vision and Pattern Recognition (CVPR). 770–778 (2016).

50. Simonyan, K., & Zisserman, A. Very deep convolutional networks for large-scale image recognition. International Journal of Computer Vision. 344–357 (2014).

51. Zeineldin, R.A., Karar, M.E., Elshaer, Z. et al. Explainability of deep neural networks for MRI analysis of brain tumors. Int J CARS 17, 1673–1683 (2022). 10.1007/s11548-022-02619-x

52. Vermeulen, C., Pagès-Gallego, M., Kester, L. et al. Ultra-fast deep-learned CNS tumour classification during surgery. Nature 622, 842–849 (2023). 10.1038/s41586-023-06615-2

53. Hollon TC, Pandian B, Adapa AR, Urias E, Save AV, Khalsa SSS, Eichberg DG, D’Amico RS, Farooq ZU, Lewis S, Petridis PD, Marie T, Shah AH, Garton HJL, Maher CO, Heth JA, McKean EL, Sullivan SE, Hervey-Jumper SL, Patil PG, Thompson BG, Sagher O, McKhann GM 2nd, Komotar RJ, Ivan ME, Snuderl M, Otten ML, Johnson TD, Sisti MB, Bruce JN, Muraszko KM, Trautman J, Freudiger CW, Canoll P, Lee H, Camelo-Piragua S, Orringer DA. Near real-time intraoperative brain tumor diagnosis using stimulated Raman histology and deep neural networks. Nat Med. 2020 Jan;26(1):52–58. doi: 10.1038/s41591-019-0715-9. Epub 2020 Jan 6. PMID: 31907460; PMCID: PMC6960329.

54. Kim JS, Han JW, Bae JB, Moon DG, Shin J, Kong JE, Lee H, Yang HW, Lim E, Kim JY, Sunwoo L, Cho SJ, Lee D, Kim I, Ha SW, Kang MJ, Suh CH, Shim WH, Kim SJ, Kim KW. Deep learning-based diagnosis of Alzheimer’s disease using brain magnetic resonance images: an empirical study. Sci Rep. 2022 Oct 26;12(1):18007. doi: 10.1038/s41598-022-22917-3. PMID: 36289390; PMCID: PMC9606115.

55. Timothy E Richardson, Seema Patel, Jonathan Serrano, Adwait Amod Sathe, Elena V Daoud, Dwight Oliver, Elizabeth A Maher, Alejandra Madrigales, Bruce E Mickey, Timothy Taxter, George Jour, Charles L White, Jack M Raisanen, Chao Xing, Matija Snuderl, Kimmo J Hatanpaa, 4Genome-Wide Analysis of Glioblastoma Patients with Unexpectedly Long Survival, Journal of Neuropathology & Experimental Neurology, Volume 78, Issue 6, June 2019, Pages 501–507, 10.1093/jnen/nlz025

56. Pizer, S. M., Amburn, E. P., Austin, J. D., Cromartie, R., Geselowitz, A., et al. Adaptive histogram equalization and its variations. Computer vision, graphics and image processing 39. 355–368 (1987).

57. Biggio B., Roli F. Wild patterns: ten years after the rise of adversarial machine learning. Pattern Recognition 84. 317–331 (2018).

58. Bortsova G, González-Gonzalo C, et al. Adversarial attack vulnerability of medical image analysis systems: Unexplored factors. Medical Image Analysis (2021).

59. Nurshazlyn Mohd Aszemi & P.D.D Dominic. Hyperparameter Optimization in Convolutional Neural Network using Genetic Algorithms. International Journal of Advanced Computer Science and Applications(IJACSA) (2019).

60. Pumperla, M. Hyperas: Simple Hyperparameter Tuning for Keras Models, IU Internationale Hochschule, Pathmind Inc. (2021).

61. Bergstra, James and Yamins, Daniel & Cox, David. Making a Science of Model Search: Hyperparameter Optimization in Hundreds of Dimensions for Vision Architectures. Proceedings of the 30th International Conference on Machine Learning. PMLR. 115—123 (2013).

62. Johny, A. K. N. D., & Nallikuzhy, D. Optimization of CNN Model With Hyper Parameter Tuning for Enhancing Sturdiness in Classification of Histopathological Images. Materials Engineering eJournal (2020).

63. Iqbal S, Qureshi AN, Ullah A, Li J, Mahmood T. Improving the Robustness and Quality of Biomedical CNN Models through Adaptive Hyperparameter Tuning. Applied Sciences (2022).

64. Banerjee, I., Bhattacharjee, K., Burns, J. L., Trivedi, H., Purkayastha, S., Seyyed-Kalantari, L., Patel, B. N., Shiradkar, R., & Gichoya, J. (2023). “Shortcuts” Causing Bias in Radiology Artificial Intelligence: Causes, Evaluation, and Mitigation. In Journal of the American College of Radiology (Vol. 20, Issue 9, pp. 842–851). Elsevier BV. 10.1016/j.jacr.2023.06.025.

65. Morgan P. McBee, Omer A. Awan, Andrew T. Colucci, Comeron W. Ghobadi et al. Deep Learning in Radiology. Academic Radiology 25. (2018)

66. Chawla, N. V., Bowyer, K. W., Hall, L. O., & Kegelmeyer, W. P.. SMOTE: Synthetic minority over-sampling technique. Journal of artificial intelligence research 16. 321–357 (2002).

67. H. Kaur & J. Rani. MRI brain image enhancement using Histogram Equalization techniques. 2016 International Conference on Wireless Communications, Signal Processing and Networking (WiSPNET). 770–773 (2016).

68. Zuiderveld & Karel J.. Contrast Limited Adaptive Histogram Equalization. Graphics gems (1994).

69. Z. Wang, A. C. Bovik, H. R. Sheikh & E. P. Simoncelli. Image Quality Assessment: From Error Visibility to Structural Similarity. IEEE Transactions on Image Processing vol. 13. 600–612 (2004)

70. J. A. Hanley and B. J. McNeil. The Meaning and Use of the Area Under a Receiver Operating Characteristic (ROC) Curve. Radiology 143.1. 29–36 (1982).

71. R. R. Selvaraju, M. Cogswell, A. Das, R. Vedantam, D. Parikh, and D. Batra. Grad-CAM: Visual Explanations from Deep Networks via Gradient-based Localization. Preprint at arXiv https://arxiv.org/abs/1610.02391 (2016).

72. Eche, T., Schwartz, L. H., Mokrane, F. Z., & Dercle, L. Toward Generalizability in the Deployment of Artificial Intelligence in Radiology: Role of Computation Stress Testing to Overcome Underspecification. Radiology. Artificial intelligence(2021).

73. Bradski, G. The OpenCV library. Dr. Dobb’s Journal of Software Tools, 120–125 (2000).

74. Geirhos, R., Jacobsen, JH., Michaelis, C. et al. Shortcut learning in deep neural networks. Nat Mach Intell 2, 665–673 (2020).

75. Chatterjee, A., Walters, R., Shafi, Z. et al. Improving the generalizability of protein-ligand binding predictions with AI-Bind. Nat Commun 14, 1989 (2023).

76. Shaeke Salman and Xiuwen Liu. Overfitting Mechanism and Avoidance in Deep Neural Networks. Preprint at arXiv https://arxiv.org/abs/1901.06566 (2019).

77. Molina D, Pérez-Beteta J, Martínez-González A, Martino J, Velasquez C, Arana E, Pérez-García VM. Lack of robustness of textural measures obtained from 3D brain tumor MRIs impose a need for standardization. PLoS One. 2017

78. Acharya, U.K., Kumar, S. Image sub-division and quadruple clipped adaptive histogram equalization (ISQCAHE) for low exposure image enhancement. Multidim Syst Sign Process 34, 25–45 (2023).

79. Zaridis, D.I., Mylona, E., Tachos, N. et al. Region-adaptive magnetic resonance image enhancement for improving CNN-based segmentation of the prostate and prostatic zones. Sci Rep 13 (2023).

80. Rahman, T. et al. Exploring the effect of image enhancement techniques on COVID-19 detection using chest X-ray images. Comput. Biol. Med. 132 (2021).

81. Otsu, Nobuyuki. A Threshold Selection Method from Gray-Level Histograms. IEEE Transactions on Systems, Man, and Cybernetics 9, 62–66. (1979).

82. Mi, H., Yuan, M., Suo, S. et al. Impact of different scanners and acquisition parameters on robustness of MR radiomics features based on women’s cervix. Sci Rep 10 (2020).

83. Deisenroth, Marc Peter, A. Aldo Faisal, and Cheng Soon Ong. Mathematics for Machine Learning. Cambridge University Press (2020).

84. Gonzalez, R.C., Woods, R.E.: Digital Image Processing, 3rd edn. Pearson Education. 129–161 (1992).

85. Rahman, S., Rahman, M.M., Abdullah-Al-Wadud, M. et al. An adaptive gamma correction for image enhancement. EURASIP Journal on Image and Video Processing (2016).

86. Joshi, V., Sukhbaatar, S., & Poczos, B. Tuning hyperparameters for deep learning: A review of recent advances. Machine Learning, 13–31 (2018).

87. Estevez, M., Benedum, C. M., Jiang et al. Considerations for the Use of Machine Learning Extracted Real-World Data to Support Evidence Generation: A Research-Centric Evaluation Framework. Cancers (2022).

88. Kingma, D. P., & Ba, J.. Adam: A method for stochastic optimization. Preprint at arXiv https://arxiv.org/abs/1412.6980 (2014).

89. Robin M. Schmidt, Frank Schneider & Philipp Hennig. Descending through a Crowded Valley - Benchmarking Deep Learning Optimizers. Preprint at arXiv https://arxiv.org/abs/2007.01547(2021).

90. Alag, M. Stochastic Gradient Descent Variants and Applications (2022).

91. Elshamy, R., Abu-Elnasr, O., Elhoseny, M. et al. Improving the efficiency of RMSProp optimizer by utilizing Nestrove in deep learning. Sci Rep 13, 8814 (2023).

92. Cheik Traoré & Edouard Pauwels. Sequential convergence of AdaGrad algorithm for smooth convex optimization. Operations Research Letters 49. 452–458 (2021).

93. Ruoqi Shen, Liyao Gao & Yi-An Ma. On Optimal Early Stopping: Over-informative versus Under-informative Parametrization. Preprint at arXiv https://arxiv.org/abs/2202.09885 (2022).

94. Xu, C., Tang, X., Lu, P. Using the Strongest Adversarial Example to Alleviate Robust Overfitting. Advanced Data Mining and Applications ADMA 2022. vol 13726 (Springer, 2022).

95. Ian J. Goodfellow, Jonathon Shlens & Christian Szegedy. Explaining and Harnessing Adversarial Examples. Preprint at arXiv https://arxiv.org/abs/1412.6572 (2015).

96. Pang, Tianyu, Min Lin, Xiao Yang, Jun Zhu & Shuicheng Yan. Robustness and Accuracy Could Be Reconcilable. Preprint at arXiv https://arxiv.org/abs/2202.10103 (2022).

