## Supplemental Table 1 for "SIENNA: Lightweight Generalizable Machine Learning Platform for Brain Tumor Diagnostics"

**Table S1: Mapped Models Datasets 2015-2024 for Figure 6a and 6b**

| | Year | Model Type | MRI datasets for TVT | Total data size | Metric: Accuracy | Trainable Parameters | Multi-classification |
|---|---|---|---|---|---|---|---|
| 1 | 2021 | Deep CNN | RIDER, REMBRAND, TCGA-LGG, Cheng et al | 2990 | 99.33 | 1,11,23,938 | Brain tumor groups overall? |
| | | | | 3950 | 92.66 | 6,28,885 | normal, glioma, meningioma, pituitary and metastatic |
| | | | | 4570 | 98.14 | 32,45,347 | Grades II, III and IV |
| 2 | 2020 | CNN | Cheng et al | 3064 | 96.56 | 33,655,939 | glioma, meningioma, pituitary |
| 3 | 2022 | CNN-LSTM | Kaggle dataset | 3264 | 92.00 | 1510273 | Normal, glioma tumor, meningioma, pituitary tumor |
| 4 | 2022 | CNN-SVM Classifier | BraTS dataset | | 96.19 | 42079040 | Glioma HGG type |
| | | | | | 95.46 | 42079040 | Glioma LGG type |
| 5 | 2023 | Transfer learning | Kaggle Brain Tumor MRI Dataset | 7023 | 98.00 (InceptionV3) | 23,851,784 | Normal, glioma, meningioma, - pituitary tumor |
| | | | | | 96.00 (VGG19) | 143,667,240 | glioma - meningioma - no tumor and pituitary |
| | | | | | 96.00 (DenseNet121) | 8,062,504 | glioma - meningioma - no tumor and pituitary |
| | | | | | 96.00 (MobileNet) | 4,253,864 | glioma - meningioma - no tumor and pituitary |
| 6 | 2021 | Deep dense inception residual network | Cheng et al | 3064 | 99.69 (Inception ResNet v2) | 9445379 | 3 class glioma, meningioma, pituitary |
| 7 | 2023 | CNN-Model | Brats-2017, Brats-2018, & Brats-2019 | | 97.85, 98.88, and 99.88 (by dataset) | 287809 | low- and high-grade gliomas |
| 8 | 2022 | CNN+ WSI and mpMRIs | Brats 2020 CPM-RadPath 2020 | | 69.90 (mpMRI-based method) | 25,000,000 | Glioma subtype peritumoral edema (ED), enhancing tumor (ET), and necrosis or non-enhancing tumor (NCR/NET) |
| 9 | 2023 | CNN hybrid | CE-MRI Figshare | 2145 | EfficientNetB0 (97.43) | 4,100,000 | glioma, meningioma, and pituitary tumors |
| | | | | | EfficientNetB1 (98.66) | 6,610,000 | glioma, meningioma, and pituitary tumors |
| | | | | | EfficientNetB2 (99.06) | 7,810,000 | glioma, meningioma, and pituitary tumors |
| | | | | | EfficientNetB3 (98.08) | 10,830,000 | glioma, meningioma, and pituitary tumors |
| | | | | | EfficientNetB4 (97.90) | 17,850,000 | glioma, meningioma, and pituitary tumors |
| 10 | 2021 | Segmentation | BRATS 2018, CE-MRI dataset | | 92.03, 91.13 and 87.26 (Dice Score) | 22,491,460 | MRI multi-modalities brain images. whole tumor, enhancing tumor, and tumor core |

| | | | | | | | |
|---|---|---|---|---|---|---|---|
| 11 | 2023 | AlexNet CNN + Multiple ML | Kaggle | 3600 | 88.75, 98.15, 86.25 and 100.00 | 62,378,344 | glioma, meningioma, and pituitary tumors |
| 12 | 2016 | Deep CNN segmentation | Brats 2013 | | InputCascadeCNN (88.00) | 802368 | brain tumor segmentation |
| | | | | | LocalCascadeCNN (88.00) | 654368 | brain tumor segmentation |
| | | | | | MFCascadeCNN (86.00) | 662513 | brain tumor segmentation |
| 13 | 2018 | CNN | Cancer Imaging Archive | 4069 | 91.16 | 213,122,435 | Low grade or High grade |
| 14 | 2022 | DACBT | Cheng et al, T. Brain tumor | 3064 + 253(Cross-validation) | ResNet50-CNN (99.10) | 25,600,000 | Meningioma, Glioma, and Pituitary |
| | | | | | VGG-16-CNN (97.88) | 138,400,000 | Meningioma, Glioma, and Pituitary |
| | | | | | Inception V3-CNN (98.02) | 138,400,000 | Meningioma, Glioma, and Pituitary |
| | | | | | DenseNet201-CNB (97.90) | 20,200,000 | Meningioma, Glioma, and Pituitary |
| | | | | | Xception-CNN (98.97) | 22,900,000 | Meningioma, Glioma, and Pituitary |
| | | | | | MobilleNet-CNN (98.56) | 4,300,000 | Meningioma, Glioma, and Pituitary |
| 15 | 2018 | CNN | Brats 2018 | | 92.98 | 281541 | Glioma ROI segmentation CNN |
| | | | | | 89.50 | 164525 | Glioma grading CNN |
| 16 | 2023 | BResNext101 _32× 8d and VGG19 | Brain-Tumor-Datasets, Kaggle Repository | 1,800 | 99.98 and 100.00 | 2325568 | Glioma and pituitary tumor |
| 17 | 2022 | Explainability of CNNs | BraTS challenges 2019 and 2021 | | 98.62 | 2097922 | Glioma classification |
| | | | | | | 25617539 | Glioma segmentation |
| 18 | 2022 | SCENIC: CNN | BraTS 2017 | | 98.30 | 168414 | Tumor detection |

Non-Brain MRI imaging application

| | Year | Model type | Metric Accuracy | Trainable Parameters | Task |
|---|---|---|---|---|---|
| 19 | 2015 | Hierarchical Feature Representation and Multimodal Fusion with Deep Learning | 95.35, 85.67, and 74.58 | 2,003,000 | Hierarchical feature reprsentation and multimodal fusion with deep learning. AD vs. healthy Normal Control (NC), MCI vs. NC, and MCI converter vs. MCI non-converter |
| 20 | 2020 | A Deep Siamese CNN | 99.05 | - | classification of dementia stages |
| 21 | 2022 | Transfer learning | VGG-16+ALFB (97.12) | 40,96,000 | Multi-classification of dementia stages in AD |
| | | | VGG-19+ALFB (98.47) | 144,000,000 | Alzheimer's Disease dementia stage classification |

| | | | | | |
|---|---|---|---|---|---|
| 22 | 2021 | CNN | 95.52 | 32,45,860 | COVID-19 severity mild, moderate, severe, and critical |
| 23 | 2020 | CNN | 99.57 | 4,08,194 | COVID-19 detection Chest X-ray normal vs disease |
| | | | 98.27 | 6,77,507 | Normal, COVID-19. pneumonia |

References


1. Irmak, E. Multi-Classification of Brain Tumor MRI Images Using Deep Convolutional Neural Network with Fully Optimized Framework. Iran J Sci Technol Trans Electr Eng 45, 1015–1036 (2021).
2. Badža, M.M.; Barjaktarović, M.Č. Classification of Brain Tumors from MRI Images Using a Convolutional Neural Network. Applied Sciences 10 (2020).
3. Ramdas Vankdothu, Mohd Abdul Hameed, Husnah Fatima, A Brain Tumor Identification and Classification Using Deep Learning based on CNN-LSTM Method, Computers and Electrical Engineering 101(2022)
4. Latif, G, Brahim, GB, Iskandar DNFA, Bashar, A, and Alghazo, J. Glioma tumors' classification using deep-neural-network-based features with SVM classifier. Diagnostics. 12 (2022)
5. Md. Monirul Islam, Prema Barua, Moshiur Rahman, Tanvir Ahammed, Laboni Akter, Jia Uddin, Transfer learning architectures with fine-tuning for brain tumor classification using magnetic resonance imaging, Healthcare Analytics 4 (2023)
6. Kokkalla, S., Kakarla, J., Venkateswarlu, I.B. et al. Three-class brain tumor classification using deep dense inception residual network. Soft Comput 25, 8721–8729 (2021).
7. H. A. Hafeez et al., "A CNN-Model to Classify Low-Grade and High-Grade Glioma From MRI Images," in IEEE Access vol. 11, pp. 46283-46296 (2023)
8. Hsu, WW., Guo, JM., Pei, L. et al. A weakly supervised deep learning-based method for glioma subtype classification using WSI and mpMRIs. Sci Rep 12 (2022).
9. Babu Vimala, B., Srinivasan, S., Mathivanan, S.K. et al. Detection and classification of brain tumor using hybrid deep learning models. Sci Rep 13 (2023).
10. Ranjbarzadeh, R., Bagherian Kasgari, A., Jafarzadeh Ghoushchi, S. et al. Brain tumor segmentation based on deep learning and an attention mechanism using MRI multi-modalities brain images. Sci Rep 11 (2021).
11. Alok Sarkar, Md. Maniruzzaman, Mohammad Ashik Alahe, Mohiuddin Ahmad, An Effective and Novel Approach for Brain Tumor Classification Using AlexNet CNN Feature Extractor and Multiple Eminent Machine Learning Classifiers in MRIs, Journal of Sensors (2023)
12. Havaei M, Davy A, Warde-Farley D, Biard A, Courville A, Bengio Y, Pal C, Jodoin PM, Larochelle H. Brain tumor segmentation with Deep Neural Networks. Med Image Anal (2016)
13. Khawaldeh, S.; Pervaiz, U.; Rafiq, A.; Alkhawaldeh, R.S. Noninvasive Grading of Glioma Tumor Using Magnetic Resonance Imaging with Convolutional Neural Networks. Applied Sciences (2018)
14. Haq, A.u., Li, J.P., Khan, S. et al. DACBT: deep learning approach for classification of brain tumors using MRI data in IoT healthcare environment. Sci Rep 12, 15331 (2022).
15. Sergio Pereira, Raphael Meier, Victor Alves, Mauricio Reyes, Carlos A. Silva, Automatic brain tumor grading from MRI data using convolutional neural networks and quality assessment, arXiv:1809.09468v1
16. S. Mohsen, A. M. Ali, E. -S. M. El-Rabaie, A. ElKaseer, S. G. Scholz and A. M. A. Hassan, Brain Tumor Classification Using Hybrid Single Image Super-Resolution Technique With ResNext101_32× 8d and VGG19 Pre-Trained Models, in IEEE Access vol. 11, 55582-55595 (2023)
17. Zeineldin, R.A., Karar, M.E., Elshaer, Z. et al. Explainability of deep neural networks for MRI analysis of brain tumors. Int J CARS 17, 1673–1683 (2022).
18. B. S. T. Naidu et al., SCENIC: An Area and Energy-Efficient CNN-based Hardware Accelerator for Discernable Classification of Brain Pathologies using MRI, 35th International Conference on VLSI Design and 2022 21st International Conference on Embedded Systems (VLSID),168-173 (2022)
19. Suk, H. I., Lee, S. W., Shen, D., & Alzheimer's Disease Neuroimaging Initiative Hierarchical feature representation and multimodal fusion with deep learning for AD/MCI diagnosis. NeuroImage 101, 569–582 (2015).
20. Mehmood, A., Maqsood, M., Bashir, M., & Shuyuan, Y. A Deep Siamese Convolution Neural Network for Multi-Class Classification of Alzheimer Disease. Brain sciences (2020).
21. Khan, R., Akbar, S., Mehmood, A., Shahid, F., Munir, K., Ilyas, N., Asif, M., & Zheng, Z. A transfer learning approach for multiclass classification of Alzheimer's disease using MRI images. Frontiers in neuroscience (2023).
22. Irmak E. COVID-19 disease severity assessment using CNN model. IET Image Process. (2021)
23. Irmak E. Implementation of convolutional neural network approach for COVID-19 disease detection. Physiological genomics, 590–601(2020)
